# Exploratory Analysis of Electronic Intensive Care Unit (eICU) Database

**DOI:** 10.1101/2020.03.29.20042028

**Authors:** Atefeh Rajabalizadeh, Javad Norouzi Nia, Nima Safaei, Mojtaba Talafidaryani, Reyhaneh Bijari, Atousa Zarindast, Fateme Fotouhi, Masoud Salehi, Mahdi Moqri

## Abstract

The monitoring of severely ill patients is a crucial procedure for every intensive care unit (ICU). By applying different data exploration methods on monitoring data, some perspective can be gained. In the present research, such monitoring data were explored in the electronic ICU (eICU) Collaborative Research Database—an ICU database collected from more than 200 hospitals and over 139,000 ICU patients across the United States. The eICU database, with its enormous quantity of remote monitoring data, could be a great resource for extracting insightful information that can help to identify potential areas of improvement in the quality of patient treatment. Important information such as patients’ vital signs, care plan documentation, stage of illness, diagnosis, and treatment is available in the database. In the present study, we explore the distribution of the data, including demographics, conditions, and diseases, and identify important patterns and relationships between features of the data. Through an exploratory analysis of the data, including the relationships between gender, ethnicity, diseases, and quality of care and mortality rates, remarkable insights were obtained. To the best of our knowledge, this is the first comprehensive exploratory analysis of the eICU database. A deep understanding of the ICU data provides the foundation for further predictive and prescriptive analyses of the data with the ultimate goal of improving ICU treatment procedures for future patients.

## 1 Introduction

Intensive care units (ICUs) mainly provide care for patients with critical vital signs [1]. With a growing population of older and sicker patients, the quality of critical care has increased [2]. Patients in the ICUs are typically monitored in order to identify any changes in the physiological status, which is deemed to be associated with deteriorating illness. This, in turn, may reveal the need for an evaluation of the treatment procedures. The monitoring systems adjacent to patients’ beds in the ICUs generate an extensive amount of data; however, a comparatively smaller proportion of patients’ monitoring results are considered as clinical documentation [3]. This massive quantity of data provides significant opportunities and simultaneously presents certain challenges in data archiving. The main difficulty in archiving includes conjunction of various information systems and management of different data types by organizing a pervasive network [4].

The electronic ICU (eICU) Collaborative Research Database provides abundant and valuable clinical information about ICU patients, who are treated as part of the Philips eICU program [5]. This database contains CSV files, each of which has a table that is separated by commas. All tables are checked to consider the safe harbor clauses of the US Health Insurance Portability and Accountability Act (HIPAA) [6]. The experiment was designed in such a manner that each patient is identified by a random numerical identifier instead of their name. This database supports many state-of-the-art techniques to extract information, such as machine learning algorithms, decision support tools, and clinical research methods [7]. By exploring this database, appropriate care of severely ill patients and adequate coverage for each patient can be achieved, and critical care telemedicine procedures can be established [8].

The eICU database, with its 31 tables, includes information about patients, their health status upon arrival and during their stay, the treatments and services provided during their visits, and their discharge status. The data were collected across several ICUs in the US, which collaborated with the program in 2014 and 2015. The recent update of the database (V2.0), which was used for the present study, was published on May 17, 2018. The “patient table” is the core table of the eICU database, and all other tables connect to it with one of the key identifiers, including “hospital,” “hospital stay,” “unit stay,” and “patient IDs”.

The database consists of information approximately 139,367 patients who stayed in an ICU at least once. In general, the data includes 166,355 hospital admissions (across 200 hospitals) and 200,859 ICU admissions. A difference between the number of patients and admissions is observed because some patients were admitted to the ICU more than once in one hospital visit or hospitalized multiple times.

Figure 1 shows the frequency of patient admissions to a hospital. This figure shows that of 139,367 unique patients, 72%, 19%, 5%, and 4% of patients were admitted to a hospital once, twice, thrice, and four times or more, respectively.

**Figure 1.**
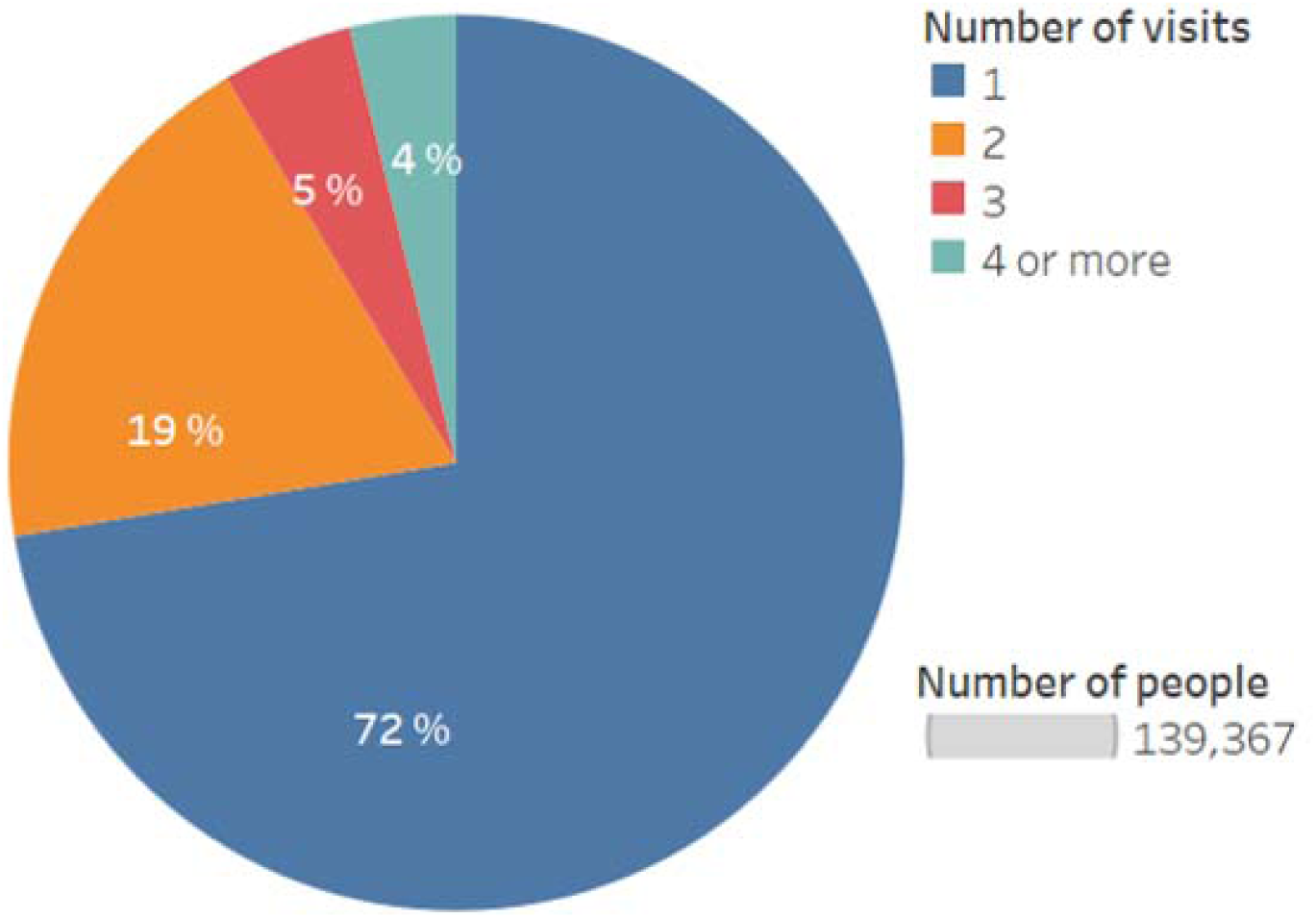
Frequency of hospital admission.

When patients leave the ICU or the hospital, the discharge status is recorded as “Expired”^1^ or “Alive”. People with “Alive” discharge status included 156,476 of the entries, whereas 9,861 of patients were discharged with the “Expired” status.

In section 2, the anomalies between mortality rates of different ages, genders, and ethnicities have been captured. In section 3, we describe the mortality rates across different ethnicity groups; we found racial bias in the eICU data. Then, we investigate it further in terms of the location of the hospital. In section 4, we explore the diseases with a high rate of mortality and frequency of death. In section 5, we present the co-presence network of diagnoses between expired patients. In section 6, we present the analysis of the patients’ length of stay at the hospital, a crucial factor in the associated costs of care and medical complications. In section 7, we examine the effects of the presence of a physician at patient discharge from the ICU on mortality rates. In section 8, we examine the effect of a physician’s presence at patient discharge from the ICU on mortality rates when a hospital stay directly followed an ICU stay. Finally, we analyze the effect of the hospital type (i.e., teaching or non-teaching) on the final results.

## 2 Demographic factors

Age, gender, and ethnicity are the demographic factors recorded in the patient table. Figure 2 shows the relationship between age distribution of patients and their status when they left the ICU. Data indicate that older patients are more likely to be referred to the ICU. The median age of ICU referral is 65 years, and mortality increases with increasing age of admission. As indicated in Figures 2 and 3, the discharge status of most of the patients is Alive.

**Figure 2.**
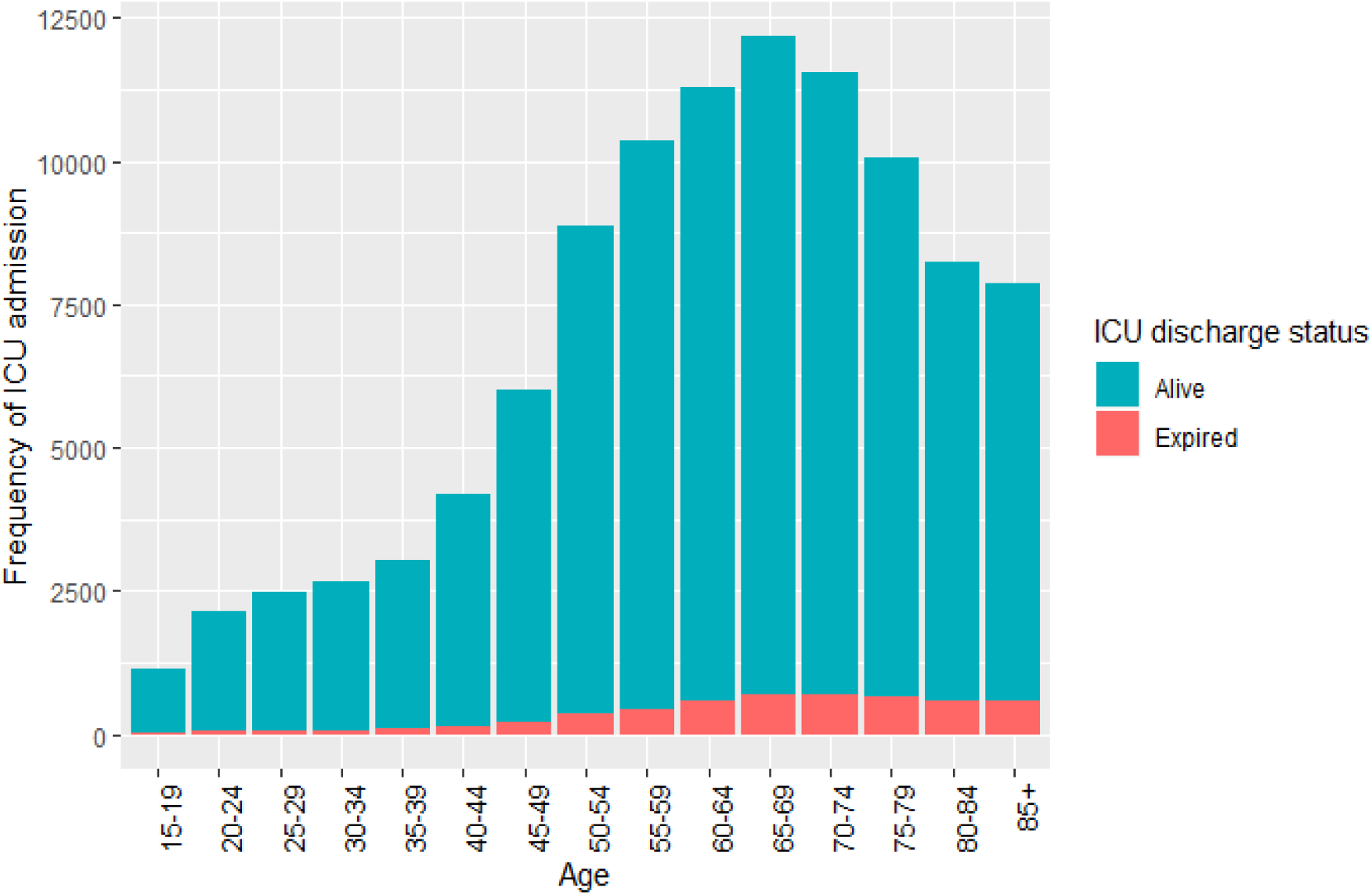
Frequency of patient admission to intensive care units among different age groups.

**Figure 3.**
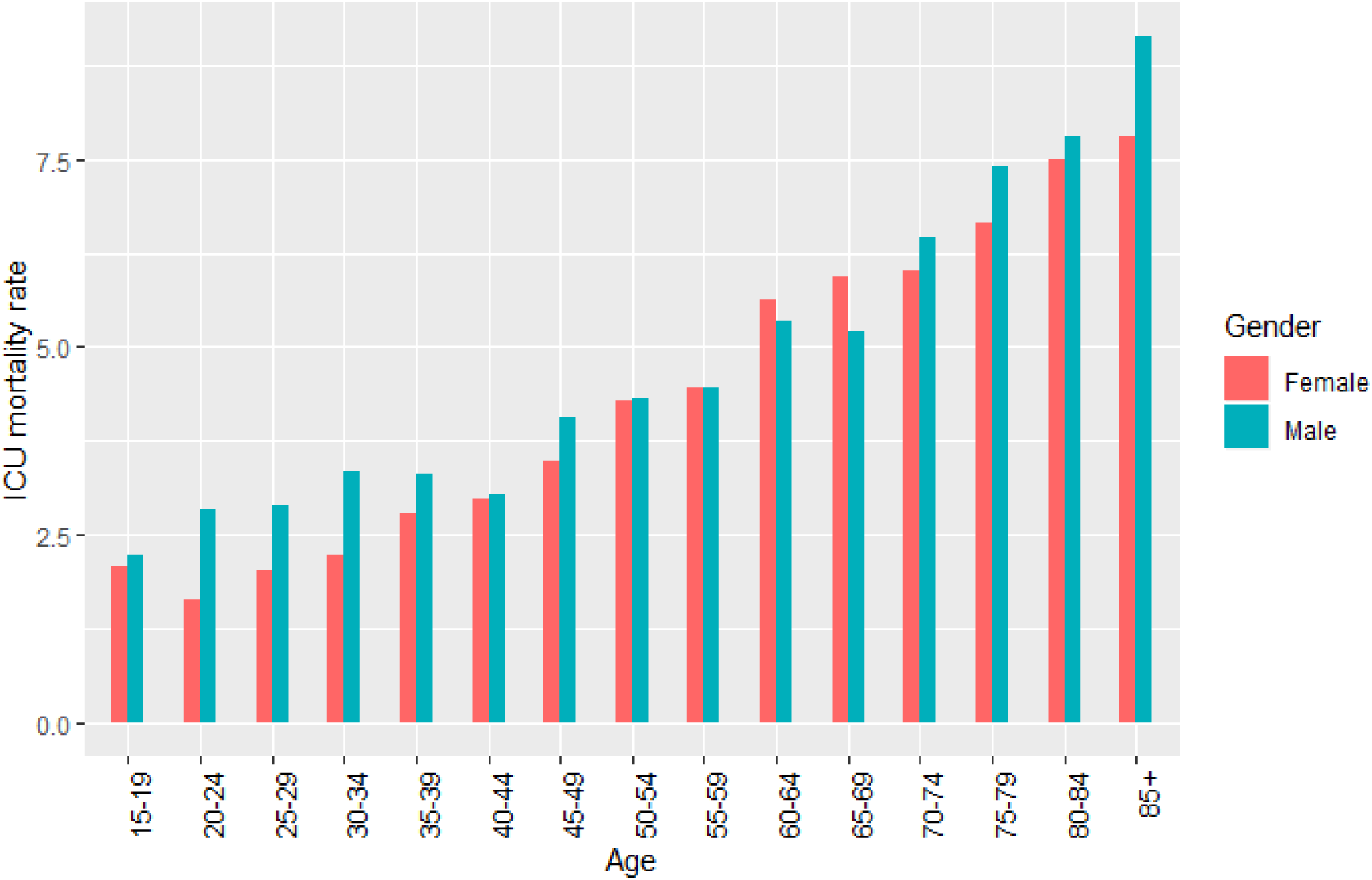
Mortality rates of intensive care unit patients according to age group and gender.

Gender data recovered from the ICU admission data indicate that 108,379 (53.95%) identified as male, 92,303 (45.95%) as female, 134 (0.06%) did not provide any information on gender, 35 (0.01%) identified as unknown, and 8 (0.003%) selected the status Other. Figure 3 presents the ICU mortality rate across different genders; the rate of mortality in men is higher than that in women in very old and very young individuals, but overall, the ICU mortality rate in both genders is almost the same (5.37% for females and 5.46% for males). The hospital mortality rate is 9.15% for females and 8.93% for males.

We also investigated the age distribution of patients who died in the ICU according to their gender (Figure 4). The median age of female patients who died in the ICU was slightly higher than that of male patients.

**Figure 4.**
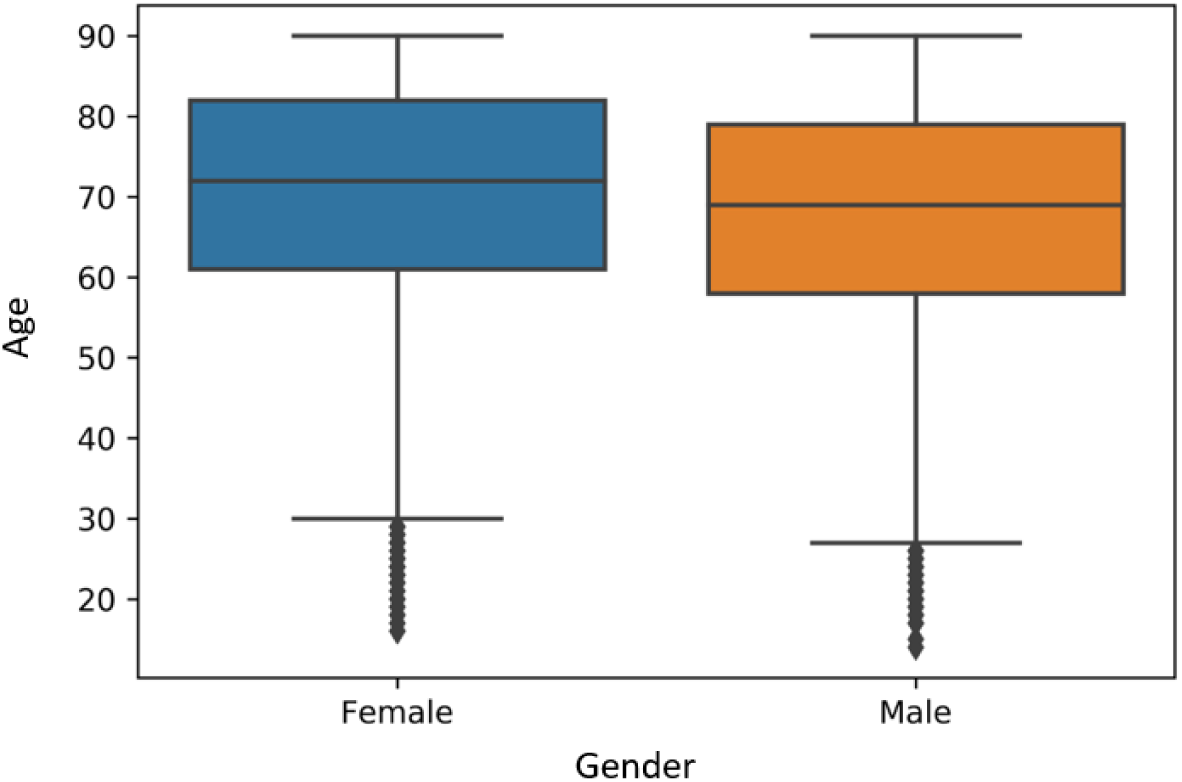
Age distribution of patients who died in intensive care units according to gender.

Ethnicity is another essential factor to study; the goal of studying ethnicity was to know the number of patients in each ethnic group. As indicated in Figure 5, Caucasians and African Americans are the two largest ethnic groups that constituted approximately 90% of the data.

**Figure 5.**
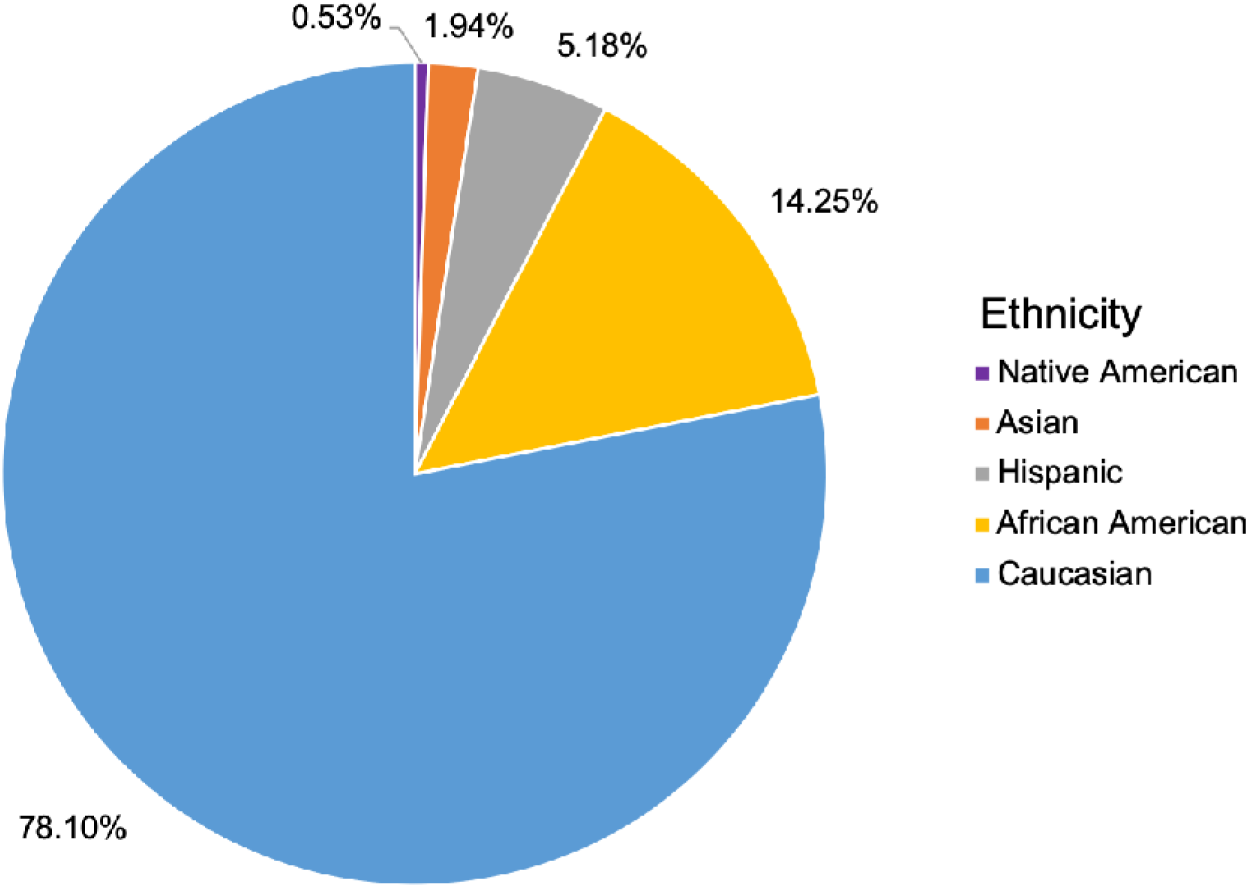
Ethnicity distribution of intensive care unit patients.

## 3 Racial bias

Figure 6 presents the comparison of the mortality rates in these two groups among different age categories. Although Caucasians have a slightly higher mortality rate (9.05%) than African Americans (8.8%) overall, a direct comparison of ICU mortality rates indicates that African Americans have a higher mortality rate in almost all age categories. This type of paradox--that one population with higher overall incidence may yet exhibit a lower incidence within each such category--is known as Simpson’s paradox [9]. The reason for this paradox is the presence of another undetected variable that plays a key role in data interpretation.

**Figure 6.**
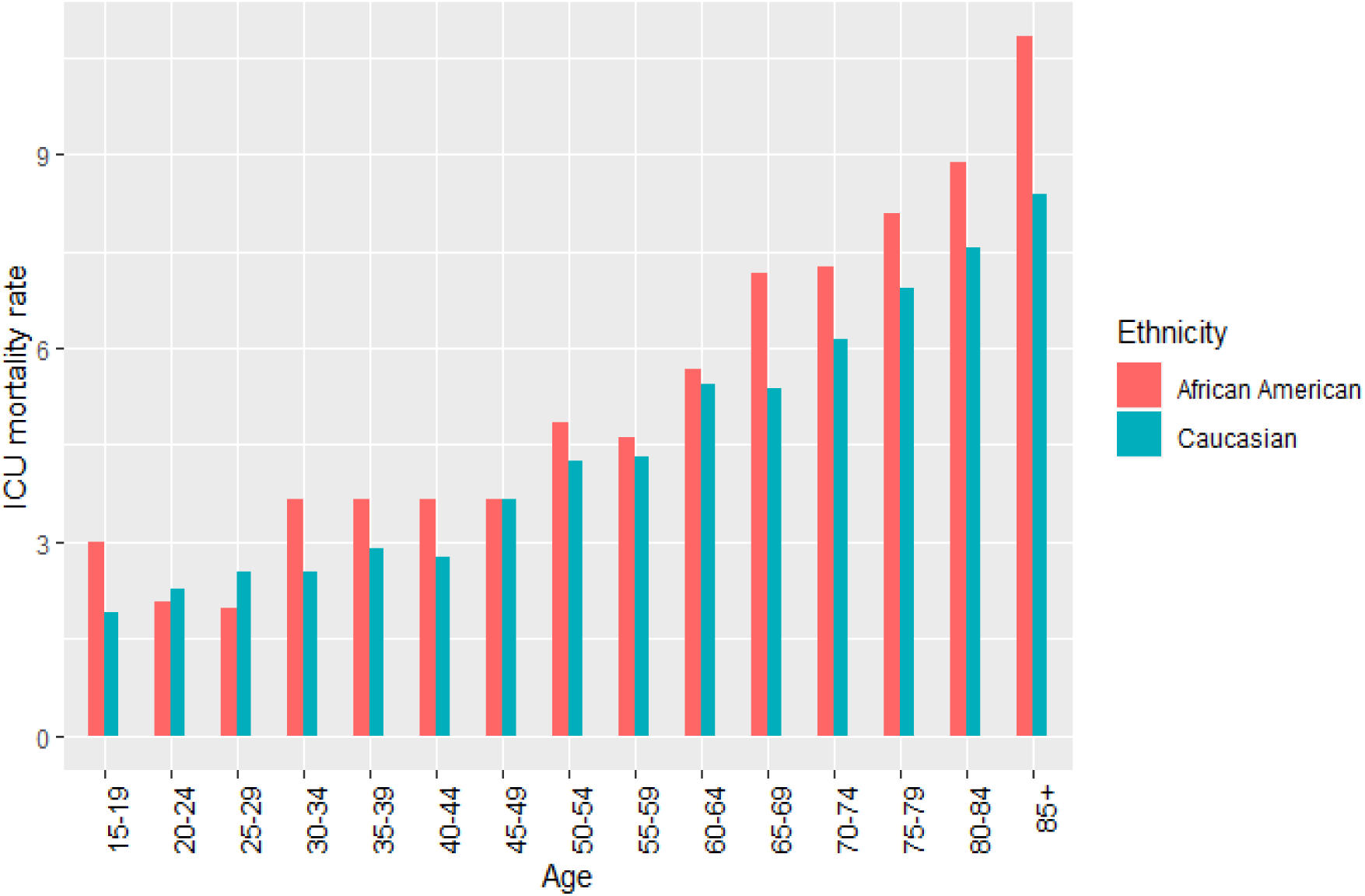
Mortality rates of intensive care unit patients per age group and ethnicity.

The higher mortality rate of Caucasian ICU patients in total may be explained by the fact that their median age is higher (as shown in Figure 7), and the frequency of death of older ICU patients is higher (Figure 7). This leads to a higher mortality rate in Caucasians in total.

**Figure 7.**
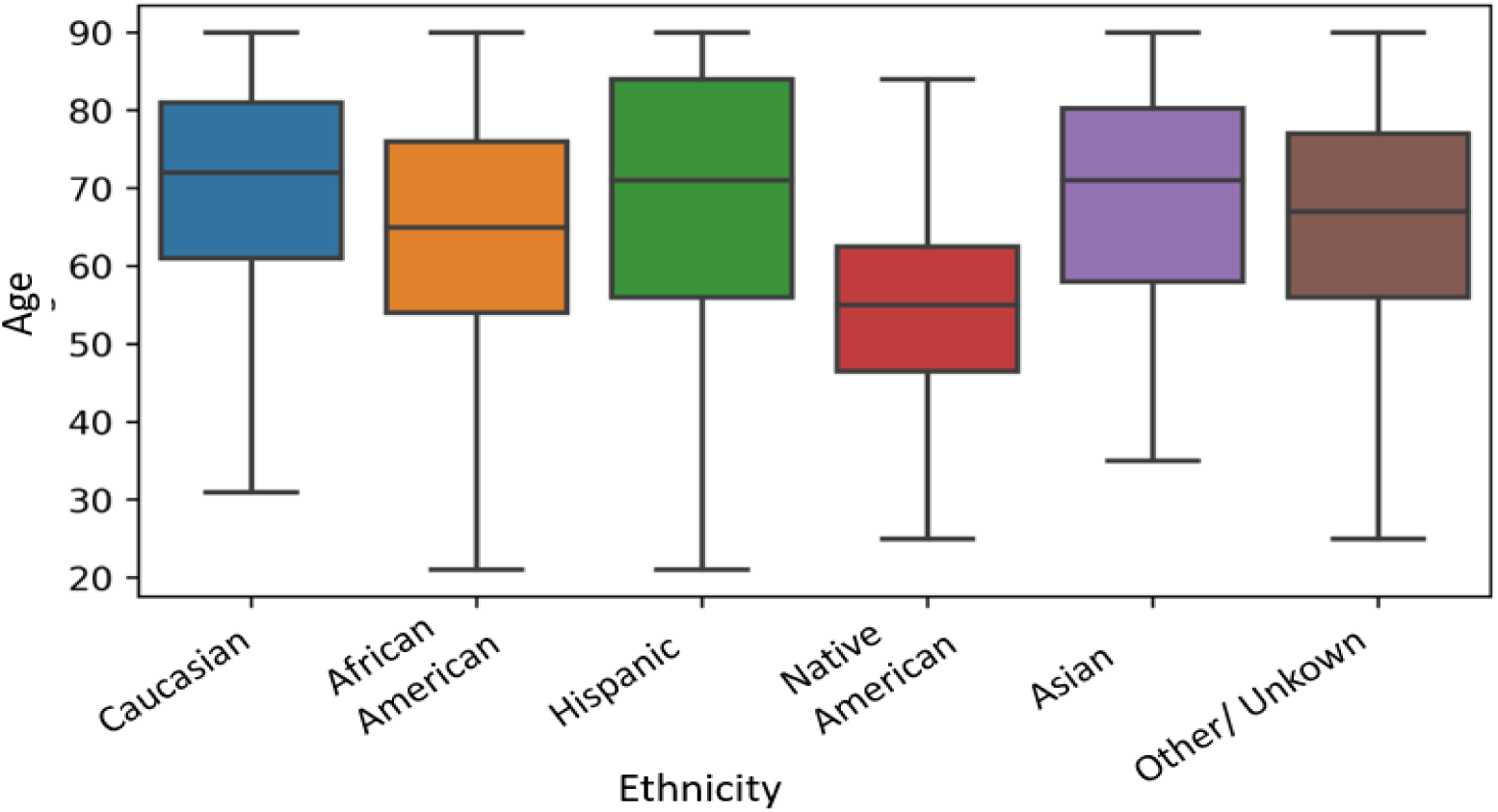
Age distribution of different ethnic groups among intensive care unit patients.

We investigated different regions to see whether the location of a hospital affected the mortality rate in different ethnic group categories (Figure 8). The majority of the monitoring data has been obtained from the South and Midwest regions of the US. In hospitals in the South, the trend of the mortality rate according to ethnicity is similar to that observed in Figure 6. In hospitals in the Midwest, however, the mortality rate of the two ethnic groups does not follow the same pattern.

**Figure 8.**
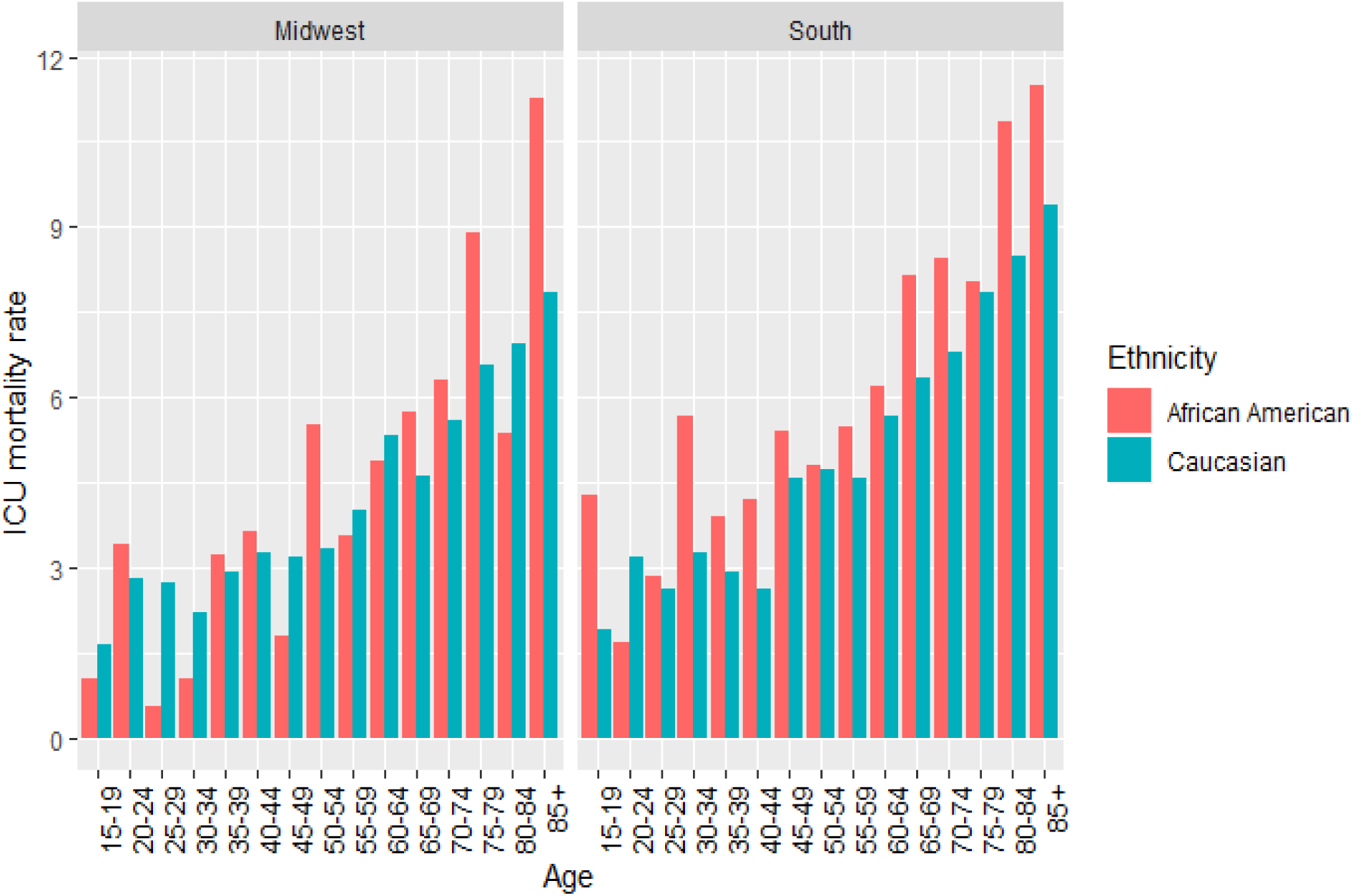
Mortality rate in intensive care unit patients according to ethnicity and age group in the Midwest and South regions of the US regions.

## 4 Admission diagnosis

Another variable of interest was the admission diagnosis for patients in an ICU unit or hospital. To classify the most important admission diagnoses, we first identified diseases with the highest frequencies and, among those, we identified the ones with the highest mortality rates.

Table 1 shows the diseases with the highest frequencies and mortality rates determined on the basis of the patients’ ICU discharge status. We considered diseases with frequencies of more than 4,500 patients and mortality rates over 10%.

**Table 1.** Most frequent diseases with the highest mortality rates based on intensive care unit discharge status.

| Disease | Patients (#) | Mortality (%) |
| --- | --- | --- |
| Sepsis, pulmonary | 8,861 | 12.05 |
| Cardiac arrest (with or without respiratory arrest; for respiratory arrest see Respiratory System) | 4,579 | 41.49 |

We also considered hospital discharge status to examine whether there were any more important diseases not found in the ICU. Apart from the diseases we previously identified, three other diseases should be considered for further investigation (Table 2).

**Table 2.** Most frequent diseases with the highest mortality rates based on hospital discharge status.

| Disease | Patients (#) | Mortality (%) |
| --- | --- | --- |
| Sepsis, pulmonary | 8,751 | 18.75 |
| CHF, congestive heart failure | 6,567 | 10.40 |
| CVA, cerebrovascular accident/stroke | 6,563 | 10.08 |
| Sepsis, renal/UTI (including bladder) | 5,212 | 11.33 |
| Cardiac arrest (with or without respiratory arrest; for respiratory arrest see Respiratory System) | 4,542 | 51.62 |

## 5 Prevalent diagnoses and their co-presence patterns among expired patients

Identifying the most prevalent diseases and their co-presence patterns in expired patients is of great importance in enhancing the treatment of future cases and lowering the mortality rate. Accordingly, we performed a text analysis for expired patients to determine whether certain diseases, either individually or in combination, predominantly led to death. To conduct the analysis, first, we aggregated the data on different diagnoses of each expired patient. Then, we identified the most prevalent diseases by performing a text-processing task based on the term frequency. The results are presented in Table 4, which ranks the diagnoses with frequencies exceeding 500 among expired patients. As this table indicates, the top 10 prevalent diseases diagnosed for expired patients are as follows: cardiovascular, pulmonary, neurological, renal, respiratory failure, gastrointestinal, endocrine, shock/hypotension, glucose metabolism, and altered mental status/pain.

It is worth noting that some of the most important ICU admission diagnoses that have been mentioned in the previous section are not included in Table 3, suggesting an inconsistency between the diagnoses documented during the ICU stay by the clinical staff and the admission diagnosis of patients.

**Table 3.** Most frequent diagnoses among expired intensive care unit patients.

| Rank | Diagnosis | Occurrences | Rank | Diagnosis | Occurrences |
| --- | --- | --- | --- | --- | --- |
| 1 | Cardiovascular | 7,853 | 27 | Acute Coronary Syndrome | 1,209 |
| 2 | Pulmonary | 4,853 | 28 | Atrial Fibrillation | 1,194 |
| 3 | Neurological | 3,957 | 29 | Congestive Heart Failure (CHF) | 1,164 |
| 4 | Renal | 3,216 | 30 | Bleeding and Red Blood Cell Disorders | 1,001 |
| 5 | Respiratory Failure | 3,052 | 31 | Anemia | 934 |
| 6 | Gastrointestinal | 2,688 | 32 | Change in Mental Status | 916 |
| 7 | Endocrine | 2,640 | 33 | Systemic/Other Infections | 890 |
| 8 | Shock/Hypotension | 2,591 | 34 | Electrolyte Imbalance | 882 |
| 9 | Glucose Metabolism | 2,281 | 35 | Stroke | 875 |
| 10 | Altered Mental Status/Pain | 2,247 | 36 | GI Bleeding/PUD | 856 |
| 11 | Disorder of Kidney | 2,233 | 37 | Hypotension | 803 |
| 12 | Chest Pain/ASHD | 2,100 | 38 | Acute Respiratory Distress | 749 |
| 13 | Acute Respiratory Failure | 2,061 | 39 | Severe | 749 |
| 14 | Arrhythmias | 1,994 | 40 | Burns/Trauma | 745 |
| 15 | Ventricular Disorders | 1,941 | 41 | Abdominal/General | 741 |
| 16 | Hypertension | 1,801 | 42 | Cardiac Surgery | 718 |
| 17 | Sepsis | 1,527 | 43 | Hyperglycemia | 711 |
| 18 | Pneumonia | 1,519 | 44 | COPD | 655 |
| 19 | Hematology | 1,496 | 45 | Coronary Artery Disease | 620 |
| 20 | Vascular Disorders | 1,460 | 46 | Chronic Kidney Disease | 610 |
| 21 | Disorders of the Airways | 1,436 | 47 | Hypoxemia | 593 |
| 22 | Acute Renal Failure | 1,420 | 48 | Toxicology | 552 |
| 23 | Infectious Diseases | 1,417 | 49 | Oncology | 518 |
| 24 | Pulmonary Infections | 1,354 | 50 | Post-GI Surgery | 514 |
| 25 | Diabetes Mellitus | 1,339 | 51 | Pain | 508 |
| 26 | Disorders of Vasculature | 1,332 | 52 | Septic Shock | 507 |

Subsequently, we investigated the co-presence patterns of prevalent diseases in expired patients by using term co-occurrence analysis, which has been an active method of research for studying disease co-occurrence in the field of network medicine [10]. The co-presence network of prevalent diagnoses among expired patients is illustrated in Figure 9. This network includes the most occurring diagnoses listed in Table 4. In Figure 9, each diagnosis is displayed by a bubble and a label, the size of which corresponds to the number of occurrences. Moreover, the distance between two nodes and the thickness of the line between them are based on the co-occurrence values of respective diagnoses. In other words, two diagnoses that frequently co-occurred among expired patients are closer together in the network and are joined by a thicker line. As shown in Figure 9, the diagnoses have been grouped into five clusters with different colors. This clustering was performed based on a network analysis technique. Each cluster consists of a group of diagnoses that often occurred together among expired patients. In fact, the network indicates that the patients who could not survive mainly suffered from one of these five major groups of diseases.

**Table 4.** Clusters of the most occurring diagnoses among expired patients.

| Cluster | Label | Diagnoses | Proportion of occurrences (%) |
| --- | --- | --- | --- |
| Red | Cardiovascular | Acute Coronary Syndrome, Arrhythmias, Atrial Fibrillation, Cardiac Surgery, Cardiovascular, Chest Pain/ ASHD, Congestive Heart Failure, COPD, Coronary Artery Disease, Diabetes Mellitus, Disorders of the Airways, Endocrine, Glucose Metabolism, Hypertension, Vascular Disorders, Ventricular Disorders | 37.07 |
| **Green** | Pulmonary | Acute Respiratory Distress, Acute Respiratory Failure, Hypotension, Hypoxemia, Infectious Diseases, Pneumonia, Pulmonary, Pulmonary Infections, Respiratory Failure, Sepsis, Septic Shock, Severe Shock/Hypotension, Systemic/Other Infections | 27.63 |
| **Blue** | Gastrointestinal | Abdominal/General, Anemia, Bleeding and Red Blood Cell Disorders, Electrolyte Imbalance, Gastrointestinal Disorders, GI Bleeding/PUD, Hematology, Hyperglycemia, Oncology, Pain, Post-GI Surgery | 13.23 |
| **Yellow** | Neurologic | Altered Mental Status/Pain, Burns/ Trauma, Change in Mental Status, Disorders of Vasculature, Neurologic, Stroke, Toxicology | 12.95 |
| **Purple** | Renal | Acute Renal Failure, Chronic Kidney Disease, Disorder of Kidney, Renal | 9.12 |

**Figure 9.**
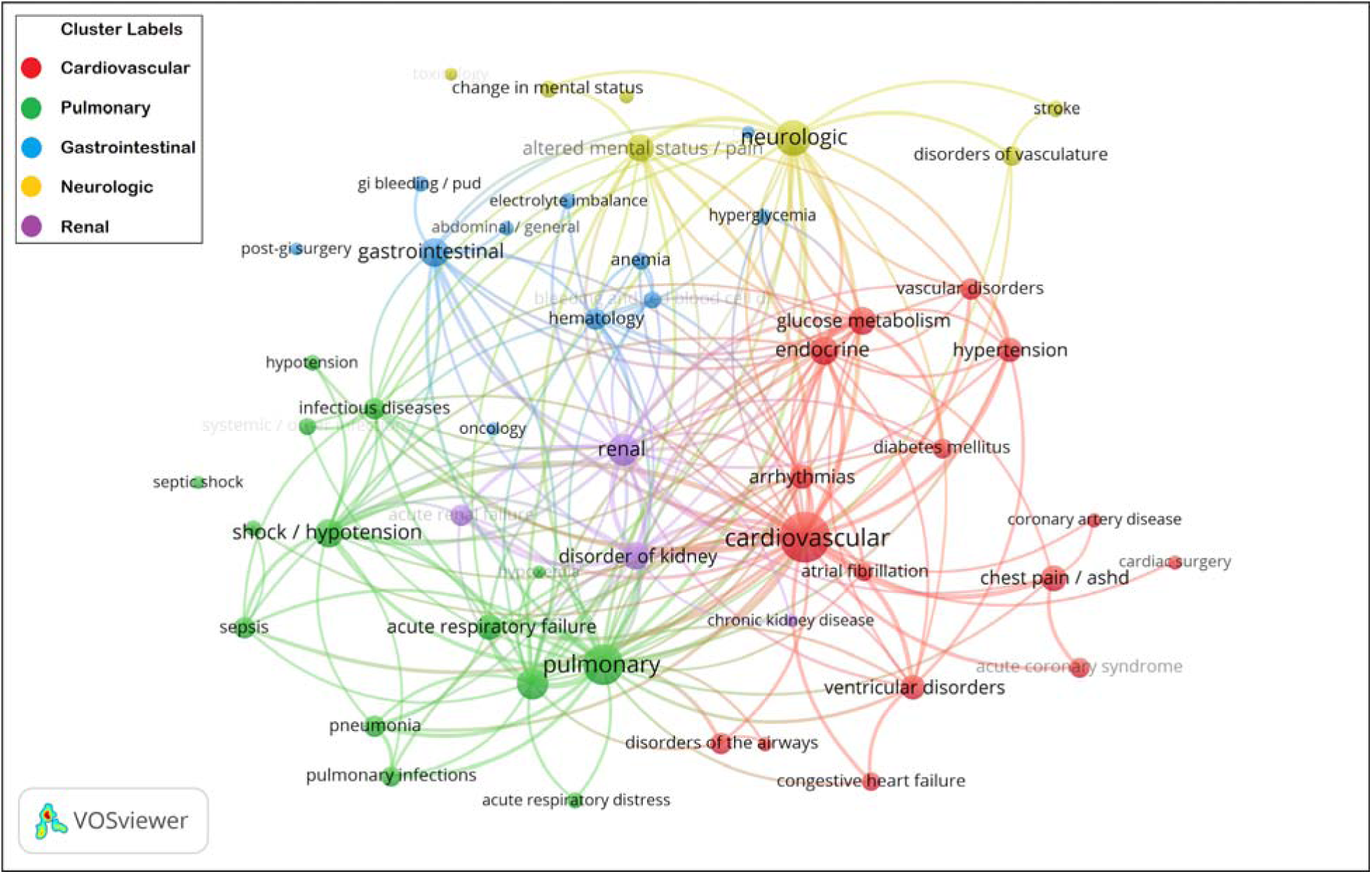
Co-presence network of prevalent diagnoses among expired patients.

Table 4 presents the respective diagnoses of each cluster. Based on the most occurring diagnosis in each cluster, we can name the red cluster as Cardiovascular, the green cluster as Pulmonary, the blue cluster as Gastrointestinal, the yellow cluster as Neurological, and the purple cluster as Renal. These results suggest that Cardiovascular, Pulmonary, Gastrointestinal, Neurological, and Renal illnesses are respectively the most prevalent groups of diseases among expired patients; that are likely more fatal and need more intensive care and attention than other illnesses.

Figure 10 depicts the density-based visualization of the co-presence network of prevalent diagnoses among expired patients. In this figure, the proximity of diagnoses represents co-occurrences. Moreover, higher occurrences of diagnoses is represented by a darker red area. Accordingly, we can recognize the more important types of diseases (e.g., cardiovascular, pulmonary, and neurological) that probably have a higher level of severity and risk of mortality. In other words, the degree of redness in an area represents the prevalence of diseases diagnosed among expired patients and can be interpreted as the mortality rate of the exhibited illnesses.

**Figure 10.**
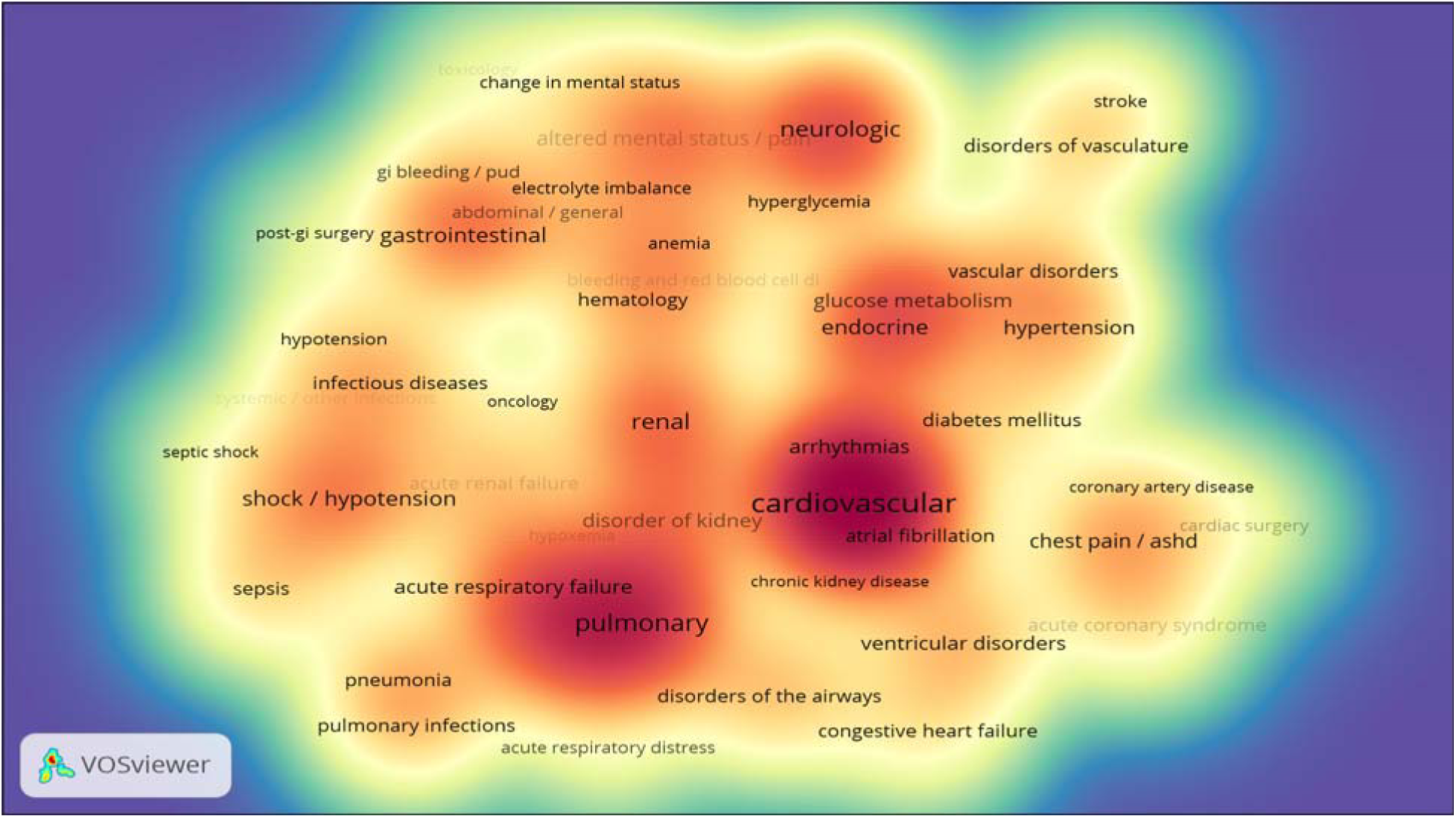
Co-presence heatmap of prevalent diagnoses among expired patients.

## 6 Length of stay in hospital

The length of stay (LOS) in hospital is a crucial metric for evaluating the efficiency of the hospital’s management system (especially the bed management). According to the Agency for Healthcare Research and Quality, the national average of LOS is 4.5 days with an average expenditure of $10,400 per day [11]. A shorter duration of hospitalization reduces the infection risk, medical side effects, hospital mortality rates, and readmission probability. It also enhances the treatment quality and increases the hospital profit; therefore, reducing the length of hospitalization (in days) has always been under careful consideration by hospital management. However, the length of time that patients spend in the ICU section is an important factor in determining the patient mortality rate.

The hospital and ICU LOS are highly correlated. One study showed that the average duration of hospitalization in a non-ICU bed increases by 1.5 days for each day that the patient spends in the ICU [12]. Therefore, the factors leading to high LOS values in the hospital and ICU should be studied together. By determining the important aspects and controlling the effective factors in predicting ICU LOS, this duration can be reduced for patients. Another study suggested that being diagnosed with cerebral infarction, middle cerebral artery, infarction of middle cerebral artery territory and myocardial infarction were the most important factors affecting the high LOS values [11]. For the eICU data, the average LOS for patients in the hospital is 8.42 days. The mode value of LOS for male and female patients is approximately 1 day and 2.5 days, respectively.

Ventilation days is another informative feature in the data that refers to the number of days a patient would be required to use a ventilator (breathing machine) in the hospital. Reducing ventilation days would reduce the risk of ventilator-associated pneumonia and would increase the chance of survival.

For this analysis, the time spent in hospital for the five most frequent diseases defined in Table 1 was investigated. Figure 11 shows the distribution of the time spent in the hospital for patients diagnosed with the five previously mentioned diseases, taking their survival status into account (“Alive” or “Expired”). Overall, across the five mentioned diseases, patients discharged with an expired status spent less time in the hospital than patients discharged with an alive status.

**Figure 11.**
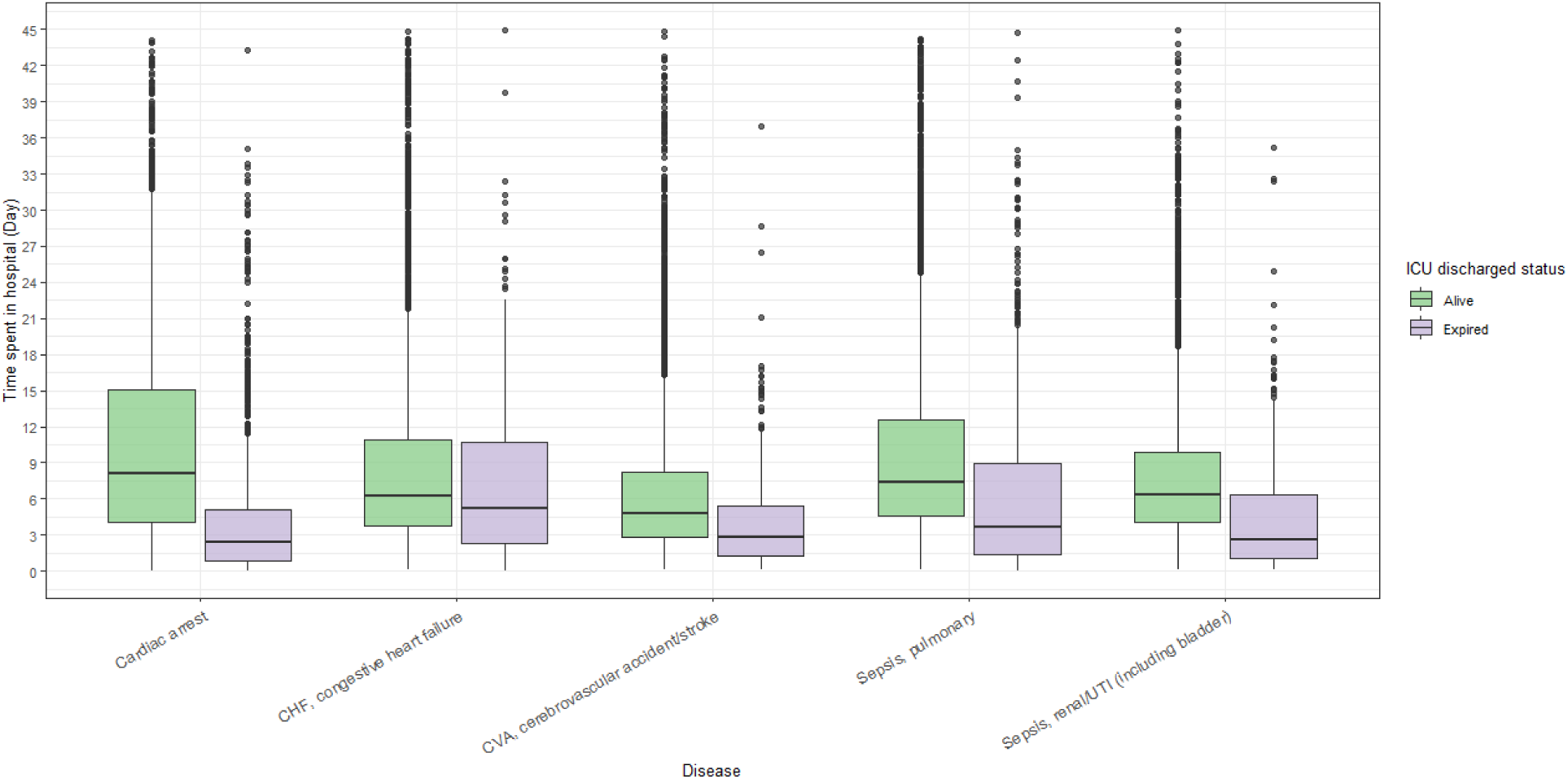
Time spent in hospital for specific diseases.

Approximately 75% of patients diagnosed with cardiac arrest who were discharged with an “Expired” status spent approximately 5 days in the hospital. The probability of being discharged alive increased for patients diagnosed with cardiac arrest who spent 6 days or more in hospital. Furthermore, 75% of patients with congestive heart failure spent approximately 10 days in the hospital regardless of their discharge status. A common pattern for cerebrovascular accident, pulmonary sepsis, and renal sepsis is that 75% of patients with expire status have died almost by day 9 (Figure 11).

We also explored the effectiveness of the predictive LOS model regarding hospital and ICU stays. Figure 12 and Figure 13 present a comparison between the predicted and actual measurements.

**Figure 12.**
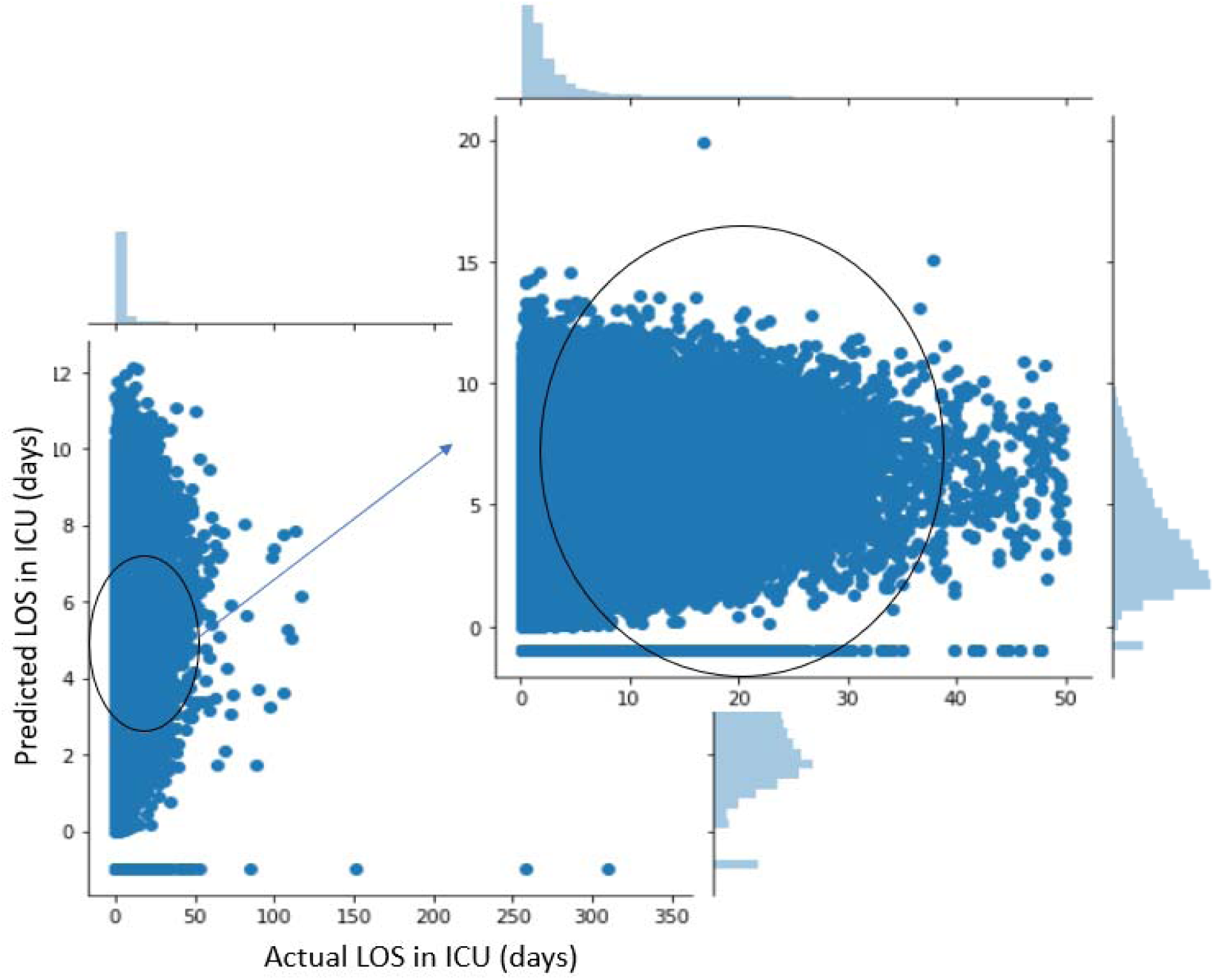
Relationship between actual length of stay (LOS) and predicted LOS among the intensive care unit (ICU) patients.

**Figure 13.**
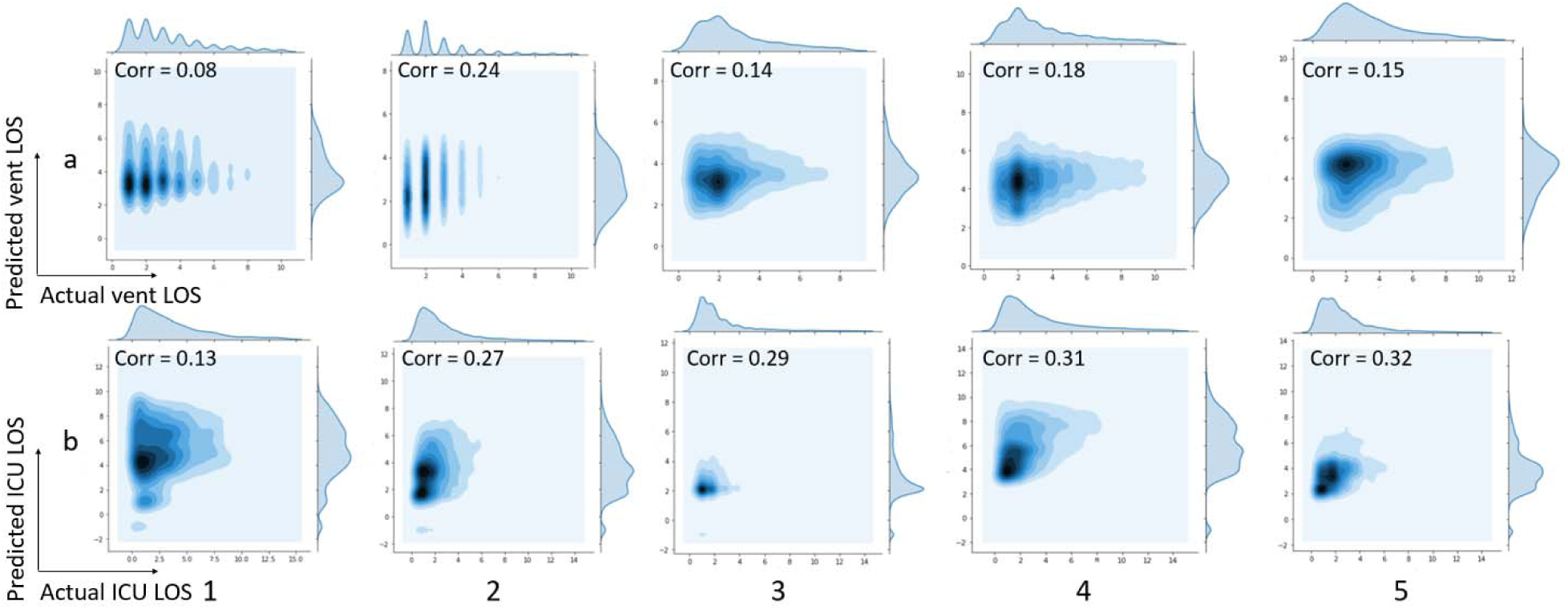
**(a) Relationship between actual and predicted ventilation days among intensive care unit (ICU) patients; 13. (b) Relationship between actual and predicted length of stay (LOS) among intensive care unit (ICU) patients**

Figure 12 shows the relationship between actual and predicted length of stay (LOS) among ICU patients. In this figure, diseases 1 to 5 are indicators of Sepsis (pulmonary), Congestive Heart Failure (CHF), Cerebrovascular Accident/Stroke (CVA), Sepsis (renal/UTI including bladder) and Cardiac Arrest respectively. The correlation coefficient of actual and predicted LOS in the ICU is 0.3. For the five major diseases defined in Table 3, Figure 13 (a) shows the kernel density estimate (KDE) and the correlation between actual and predicted ventilation days in the ICU. It is important to keep the LOS as short as possible for each patient because ventilators can have numerous side effects on patients’ respiratory systems. Figure 13 (b) shows the KDE and the correlation between the actual and predicted values of LOS in the ICU. As more than 97% of actual LOS values were lower than 14 days, we excluded LOS data that exceeded 14 days, to reduce the variance. The darker regions in the plots indicate the dense overlapping of the actual and predicted LOS values. As the graphs show, the highest density is due to the actual LOS of up to 3 days in the ICU. The predicted values are higher than the actual values, approximately double in all the represented major diseases except for congestive heart failure. We also observed a low correlation between predicted and actual values.

Table 5 shows the root mean square error (RMSE) of predicted hospital LOS, ICU LOS, and length of ventilation period. Also, the information regarding the mean and range of actual values are provided in the table to provide a basis for evaluating the results. Given the range of the dependent variables, the calculated RMSE are relatively high in all of the disease groups. Improving the accuracy for predicting these variables would help in decreasing the death and readmission rates.

**Table 5.**
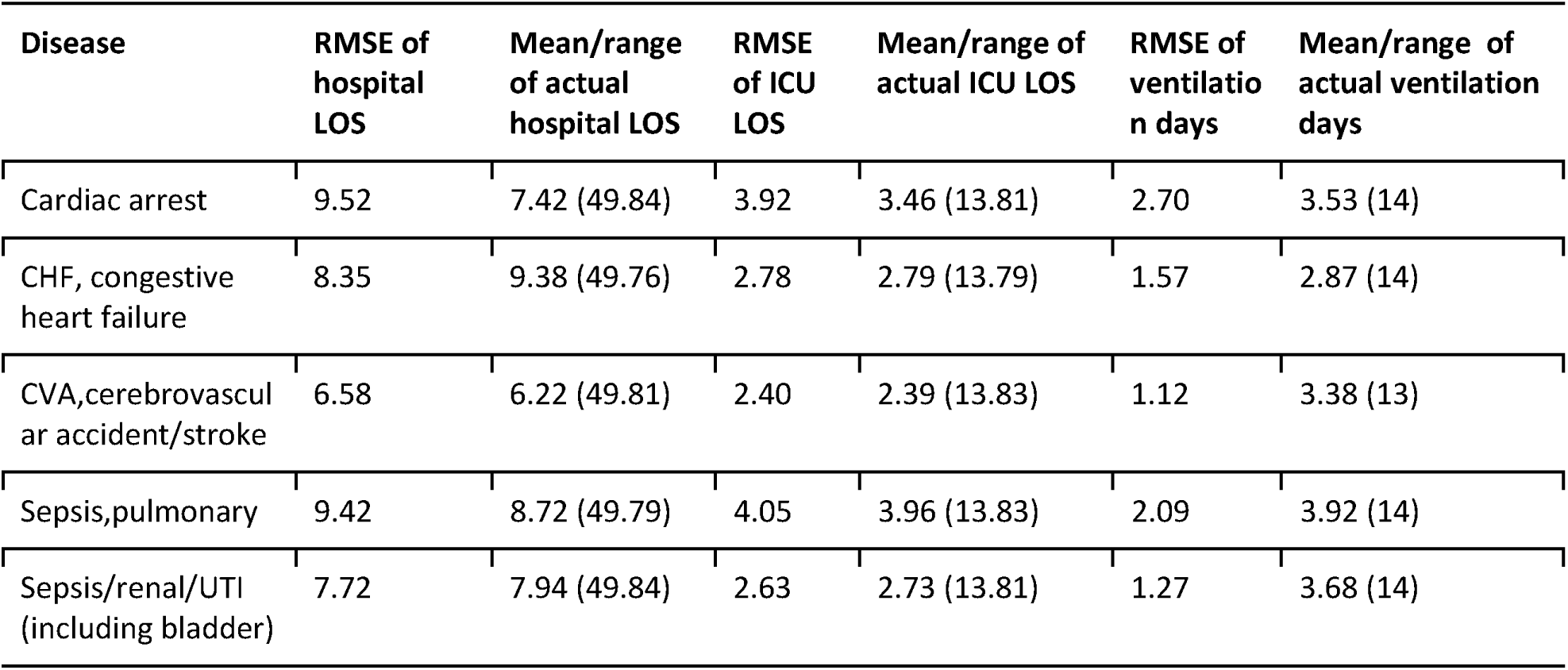
Root mean square error (RMSE) of predicted hospital and ICU LOS and predicted ventilation days.

| Disease | RMSE of hospital LOS | Mean/range of actual hospital LOS | RMSE of ICU LOS | Mean/range of actual ICU LOS | RMSE of ventilation days | Mean/range of actual ventilation days |
| --- | --- | --- | --- | --- | --- | --- |
| Cardiac arrest | 9.52 | 7.42 (49.84) | 3.92 | 3.46 (13.81) | 2.70 | 3.53 (14) |
| CHF, congestive heart failure | 8.35 | 9.38 (49.76) | 2.78 | 2.79 (13.79) | 1.57 | 2.87 (14) |
| CVA,cerebrovascular accident/stroke | 6.58 | 6.22 (49.81) | 2.40 | 2.39 (13.83) | 1.12 | 3.38 (13) |
| Sepsis,pulmonary | 9.42 | 8.72 (49.79) | 4.05 | 3.96 (13.83) | 2.09 | 3.92 (14) |
| Sepsis/renal/UTI (including bladder) | 7.72 | 7.94 (49.84) | 2.63 | 2.73 (13.81) | 1.27 | 3.68 (14) |

## 7 Effect of presence of an active physician at the time of patient’s discharge

Figure 14 clearly indicates a higher mortality rate among patients who did not have an active physician present at the time of discharge. The difference in the mortality rates of patients owing to the presence or absence of an active physician was over 4%, and it is consistent between males and females.

**Figure 14.**
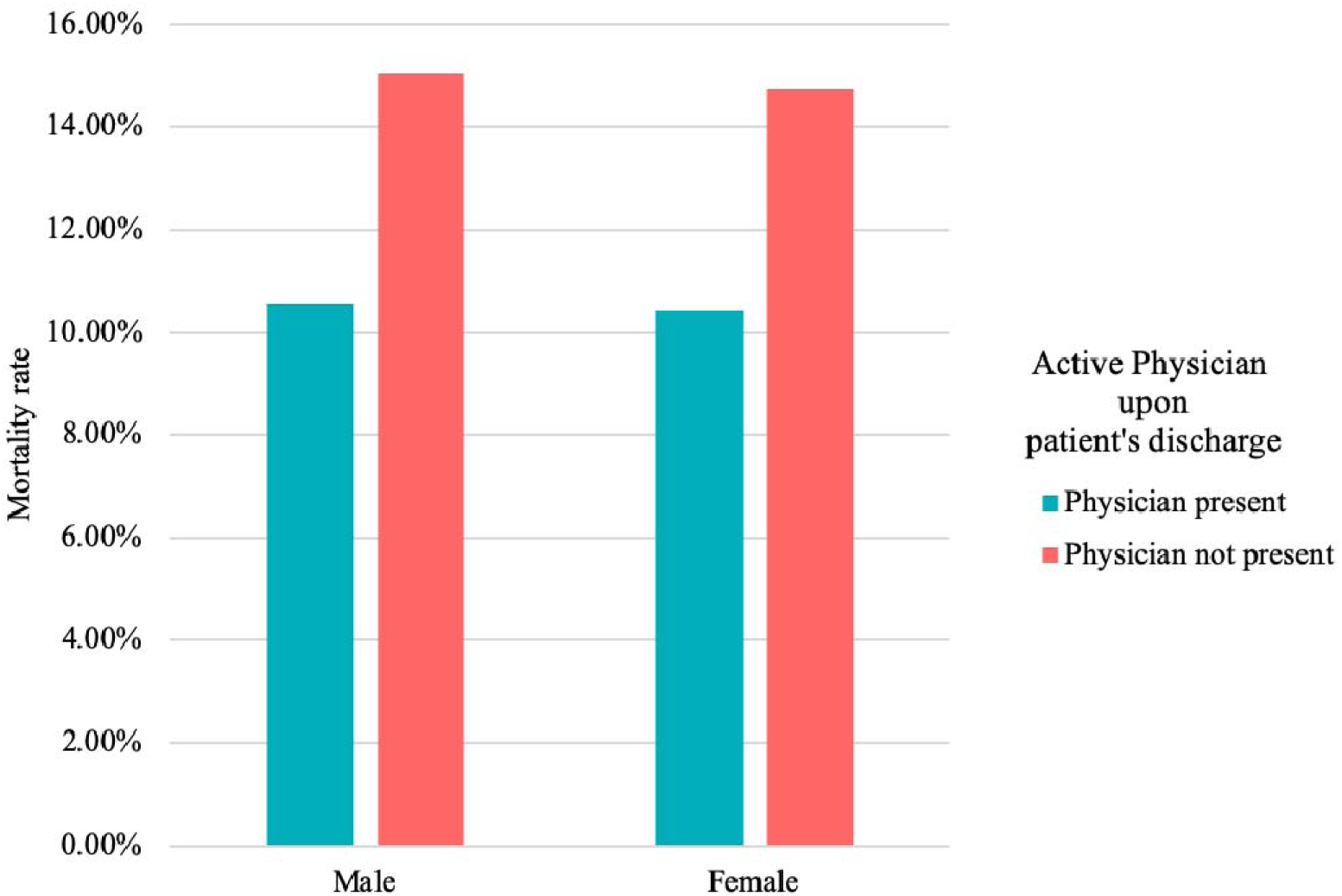
Effect of the presence of an active physician at patient discharge on the mortality rate of different genders.

Figure 15 shows the remarkable effect of meeting an active physician before discharge among different age groups. Remarkably, a rising trend is evident as we move towards older age groups.

**Figure 15.**
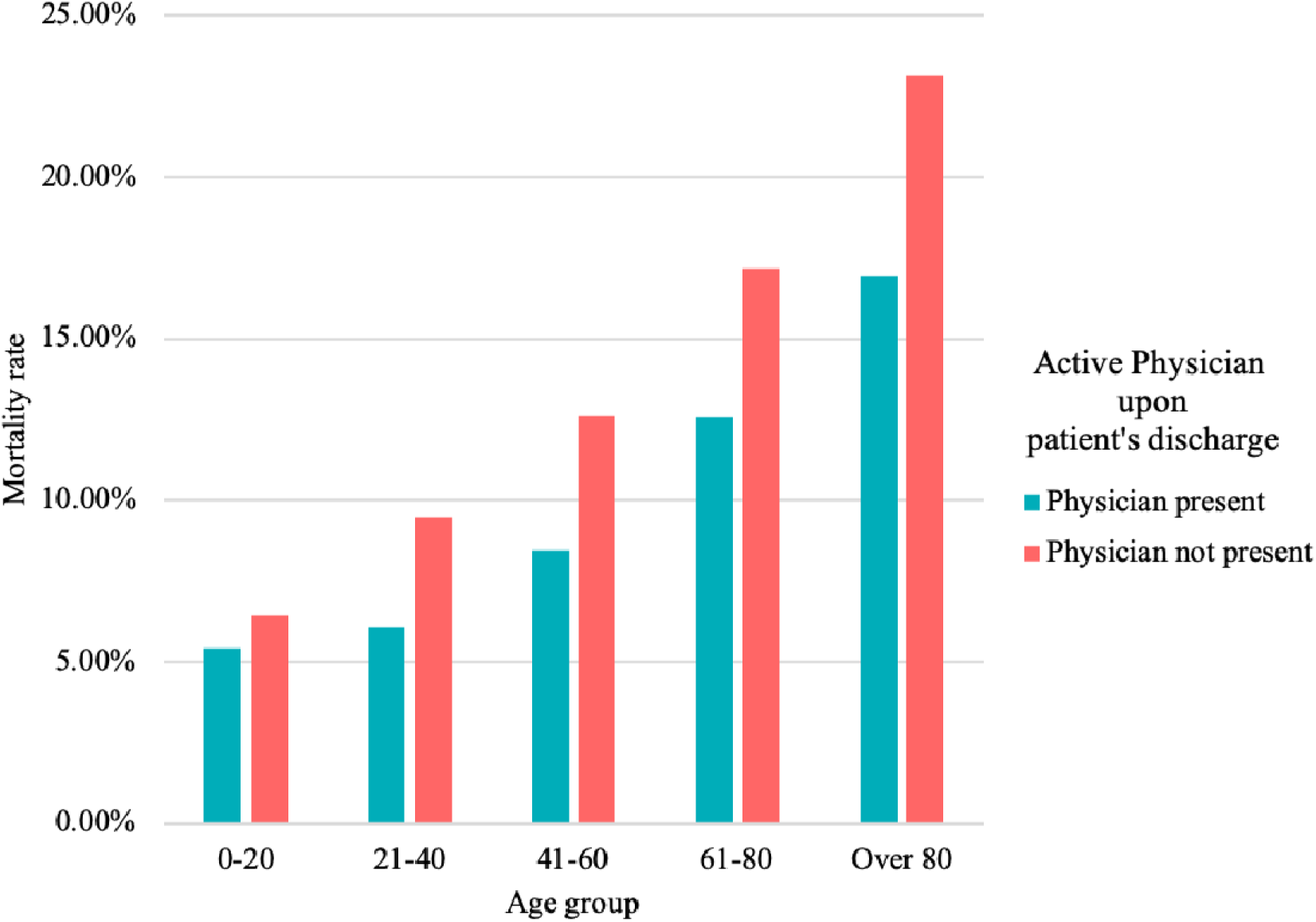
Effect of the presence of an active physician at patient discharge on the mortality rate of different age groups.

## 8 Time of discharge from hospital and intensive care unit

Figures 16a and 16b show the distribution of the time of discharge from the hospital and ICU for all patients in the database. As the graphs represent, there was a higher number of patients discharged at the start and end of the day. The highest load of discharges happened between 7:00 p.m. and 9:00 p.m.; on the other hand, the lowest load happened between 8:00 a.m. and noon.

**Figure 16.**
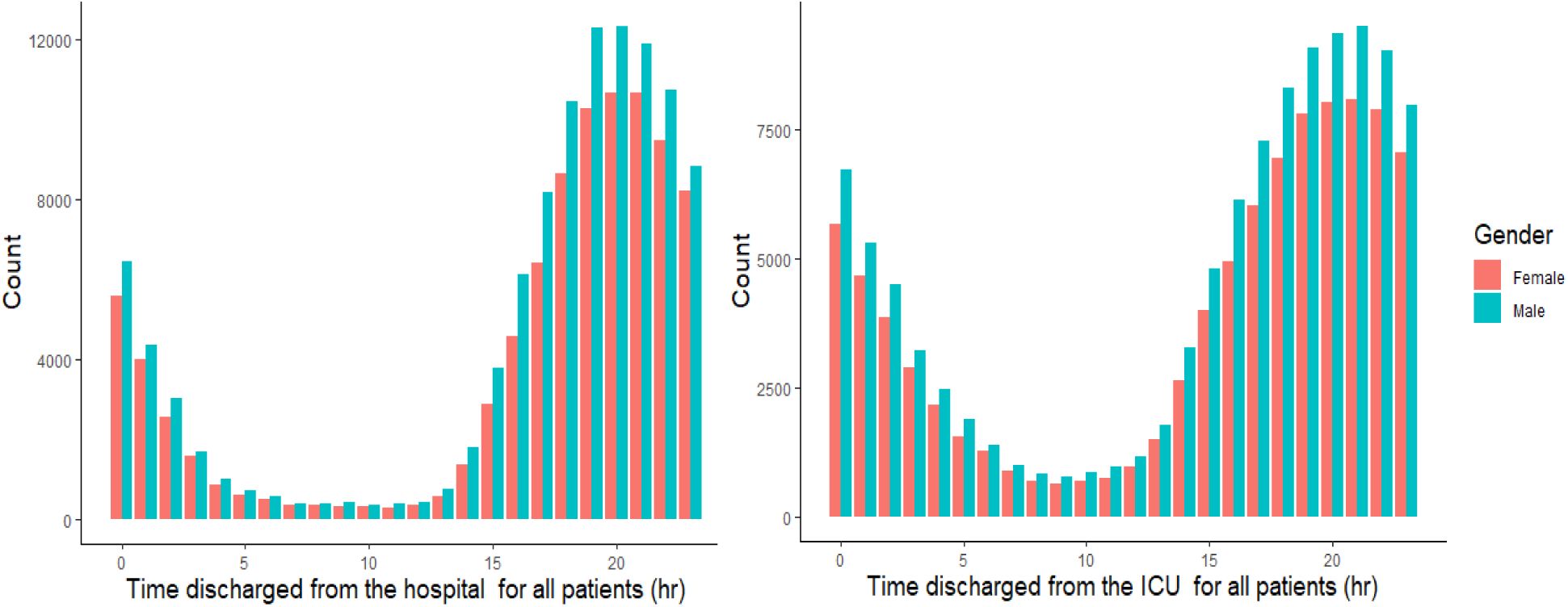
Distribution of time of discharge over 24 h of a day for all patients (a) from the hospital (b) from the intensive care unit (ICU)

Figures 17a and 17b show that the trend is rather different for expired patients. The distribution of the time of discharge from the hospital for expired patients (patients died in the hospital) indicates that the difference between different hours is less extreme, and the trend of the distribution of the time of discharge from the ICU for expired patients is slightly smoother.

**Figure 17.**
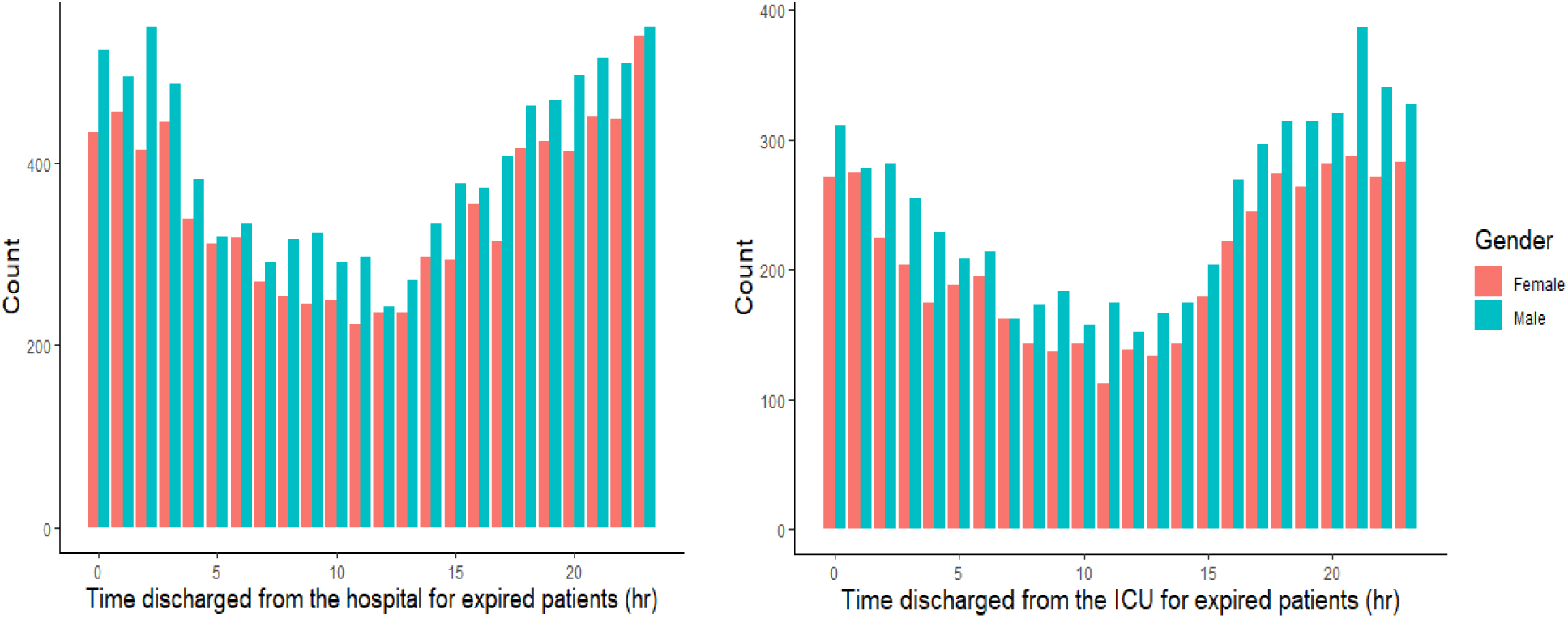
Distribution of the time of discharge over 24 h of a day for expired patients (a) from the hospital (b) from the intensive care unit (ICU)

As observed from the death rate for all patients in figure 17, it can be noted that, in general, the number of deaths is higher at night. However, this finding warrants more investigation.

The findings reported in Section 7 suggest that the presence of an active physician at patient discharge plays an important role in mortality rate. Figures 18a and 18b show the relationship between the number of expired patients from ICU and hospital, their time of discharge from the ICU at different hours of the day, and the presence or absence of an active physician at the time of their discharge from the ICU. Figure 18a suggests that the total number of expired patients did not differ significantly; however, Figure 18b indicates that discharge from the ICU in absence of an active physician was associated with considerably more deaths than discharge in the presence of an available physician. This became more critical when discharge happened at nighttime. Previous studies have also reported an association between nighttime ICU discharge and increased risk of hospital mortality [13, 14].

**Figure 18.**
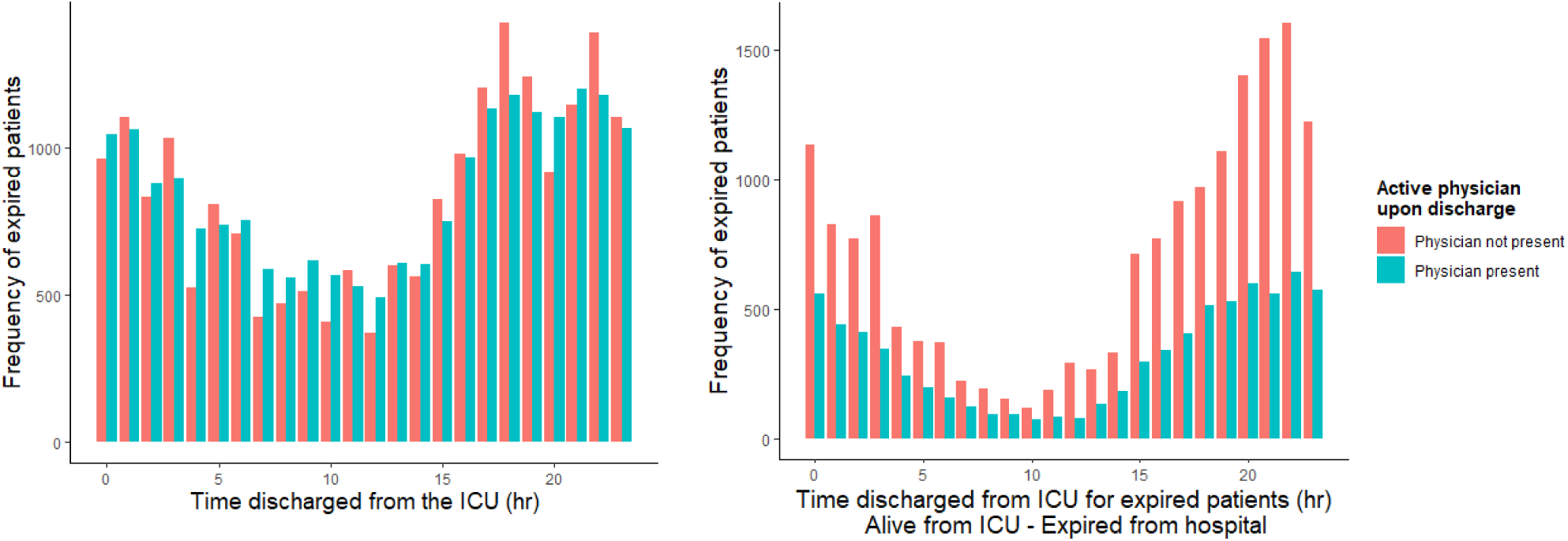
Distribution of time of discharge of expired patients from ICU (a) and hospital (b) according to active physician availability.

## 9 Hospital type

We compared the mortality rate in teaching and non-teaching hospitals and found that teaching hospitals had a higher mortality rate (Figure 19). More importantly, the mortality rate of teaching hospitals in 2015 shows an increasing trend in comparison with that in 2014.

**Figure 19.**
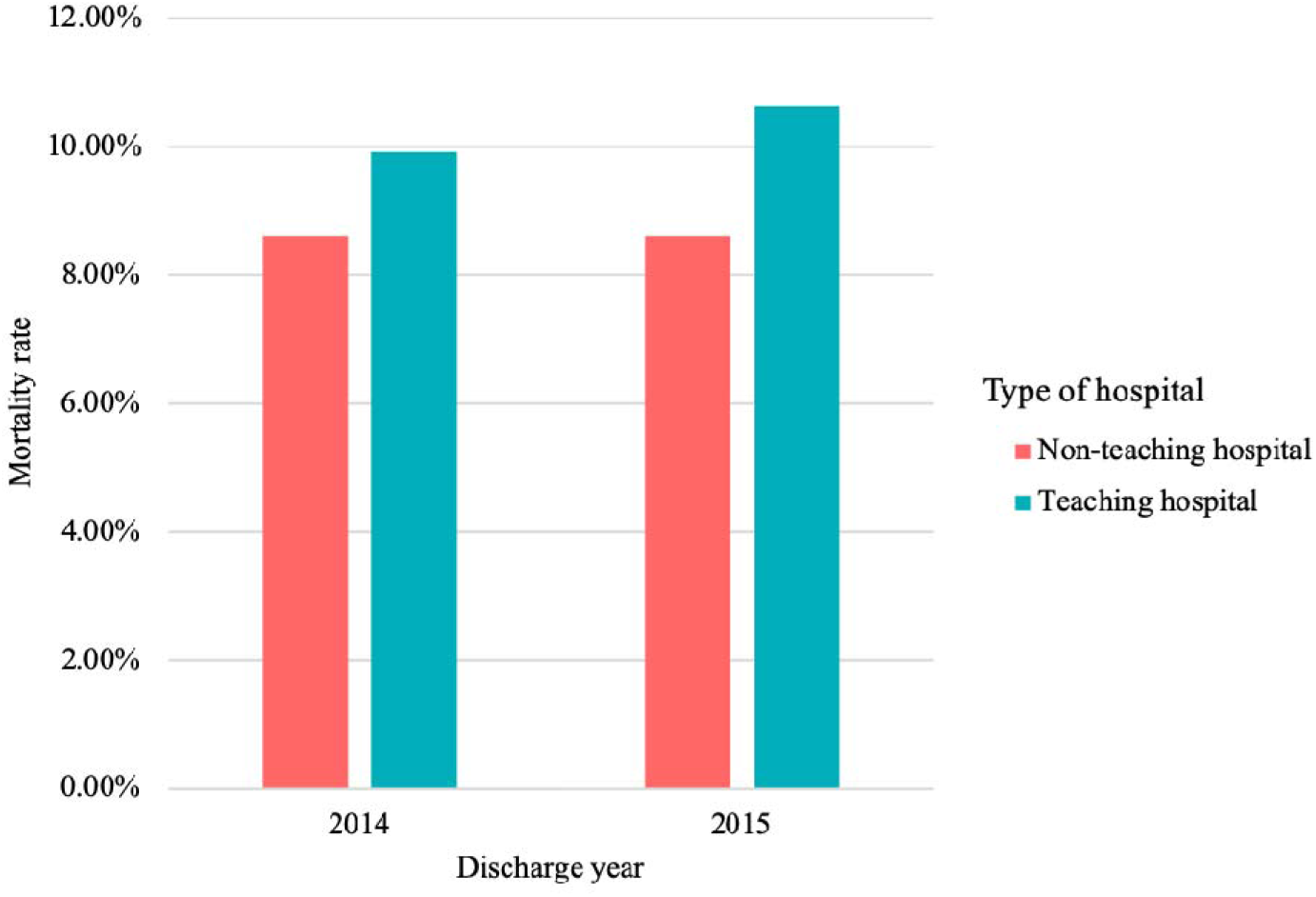
Mortality rate in teaching and non-teaching hospitals in 2014 and 2015.

However, the higher mortality rate is not consistent over every stay type such as admission, readmission, transfer, and step-down/other (Figure 20). For example, among readmitted patients, the mortality rate in teaching hospitals is lower than that in non-teaching hospitals.

**Figure 20.**
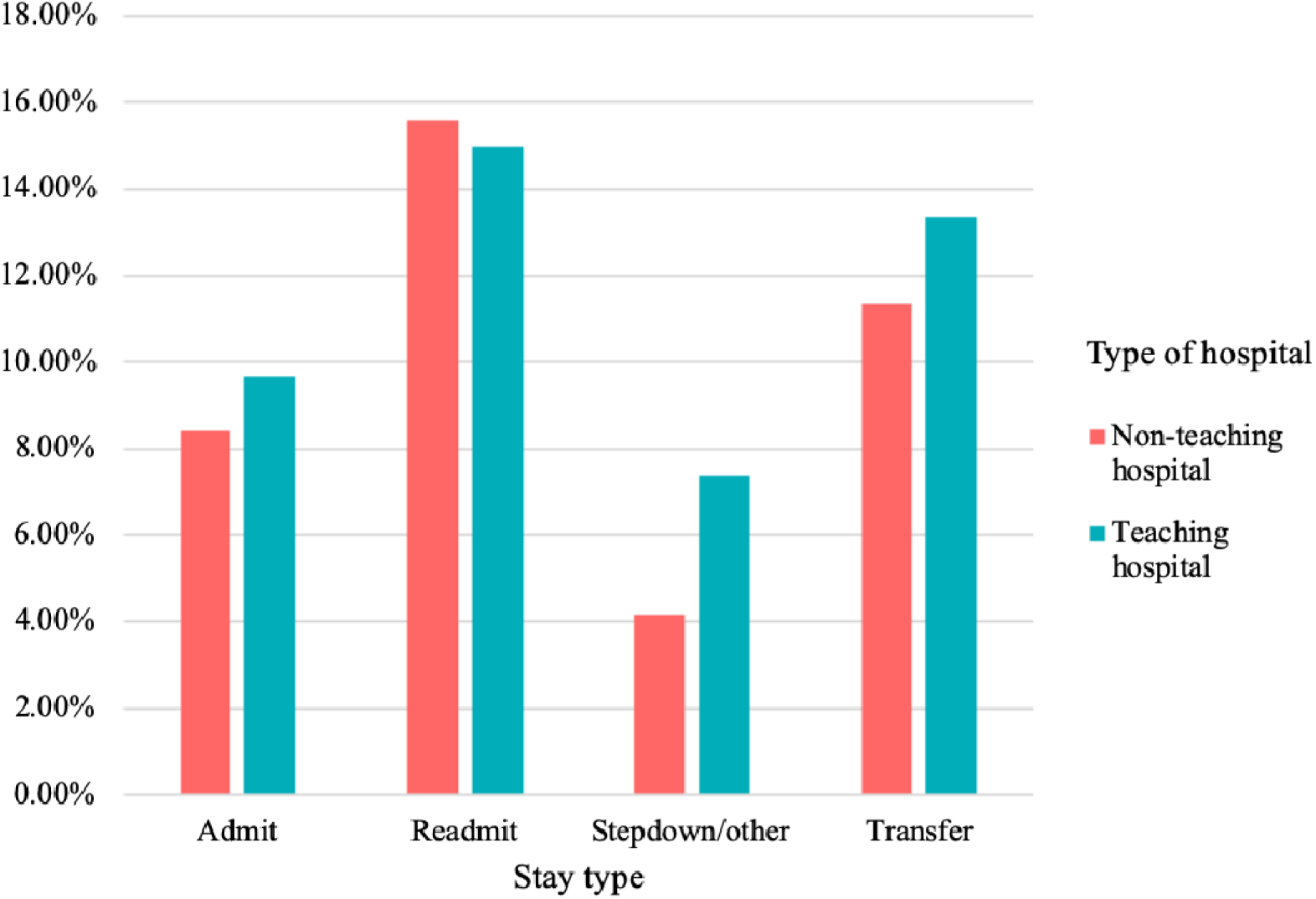
Mortality rate in teaching and non-teaching hospitals based on stay type.

Based on our findings, we expected to observe a higher mortality rate in teaching hospitals. To investigate whether this phenomenon is consistent in every ward, we analyzed the mortality rate in multiple wards (Figure 21). In many types of ICUs, the mortality rates agreed with the findings presented in Figure 20, but in neurological ICUs, the mortality rates in teaching and non-teaching hospitals were almost the same. Remarkably, in cardiothoracic ICUs, the mortality rate in teaching hospitals was distinctly lower.

**Figure 21.**
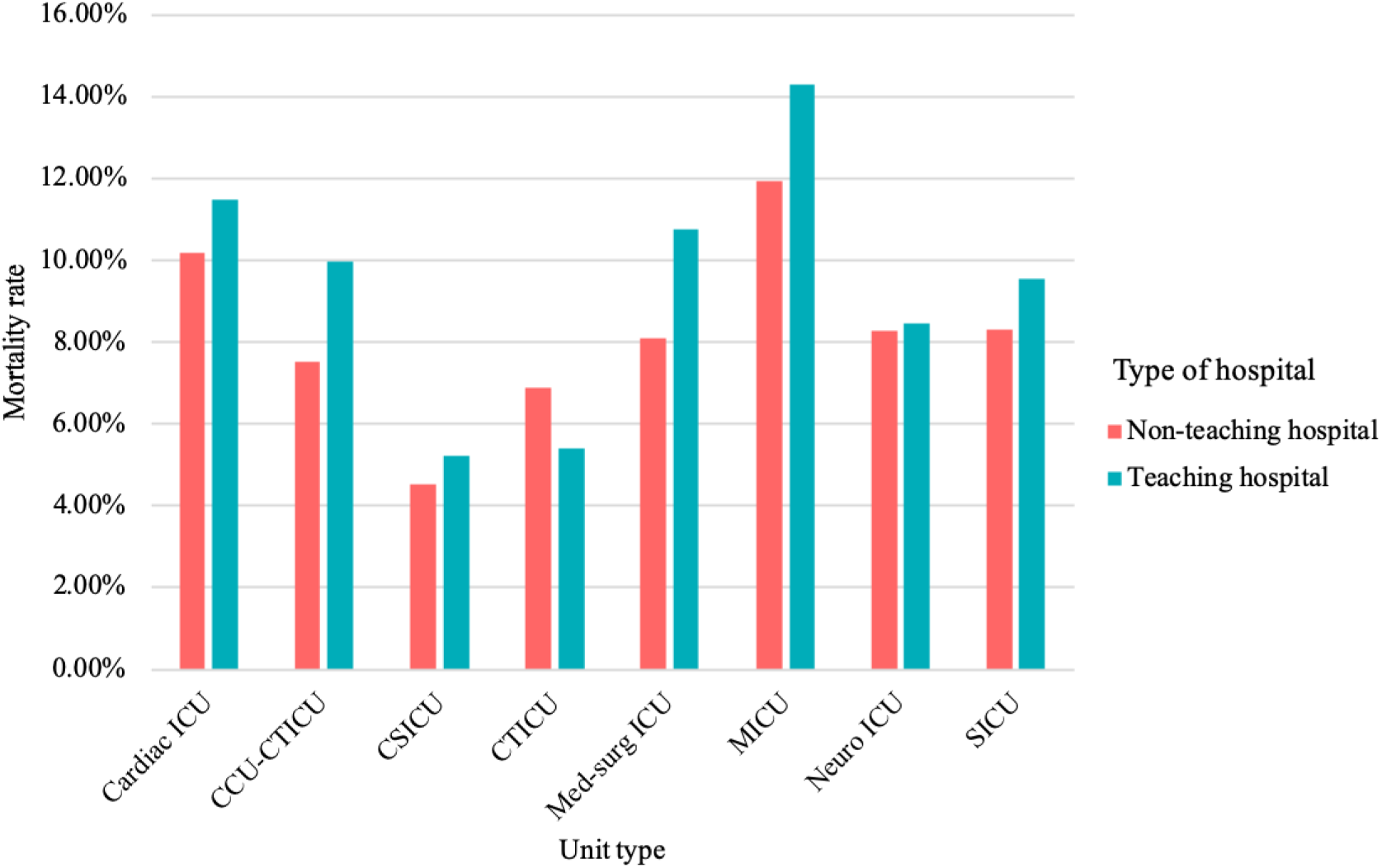
Mortality rate in teaching and non-teaching hospitals based on unit type^2^.

## 10 Discussion

In the present study, we conducted exploratory data analysis with the aim of gaining insights into the eICU database. We used different graphical representations and summary statistics to provide a better understanding of the data, which further could serve as a tool for future hypothesis generation and model development [15, 16] for critical care patients.

The analyses performed in this study could be divided into two main groups. The first group focused on patient characteristics and status, and the second group focused on quality of care and hospital characteristics. The first group includes the analysis of the patient demographic information, status, disease(s), and diagnoses. The second group includes the analysis of the length of stay, presence of an active physician at patient discharge, time of admission and discharge, and type of hospital. These two groups are potentially the most critical determinants for mortality rates in patients [17].

Based on patient-related analyses, we found that the ICU discharge status of a vast majority of the patients was “Alive”, and the rate of mortality in men and women was about the same.

One of the most important results is regarding racial bias as we found that mortality rates of African Americans were higher than that of Caucasians in almost every age category. The reason for the higher mortality rates of African Americans in most age groups is a very noteworthy topic of research and can be examined by controlling covariates. Identifying the reason for this bias can lead to new legislation towards this ethnic group.

We identified five important diseases diagnosed upon admission based on their frequency and mortality rates. Furthermore, we performed a textual analysis of the patient diagnosis data and identified the most prevalent diagnoses among expired patients. A co-presence network of diagnoses was also generated, which could be used to demonstrate the most highly co-present symptoms in expired patients.

We further investigated hospital-related data including patient LOS. The RMSE of the predicted hospital LOS across the discussed disease groups was found to be 9. The RMSE values of the predicted ICU LOS and number of ventilation days across the discussed disease groups were lower (approximately 2). Improving the accuracy of the predicted LOS is essential owing to the high correlation between longer hospital stays and higher mortality rates. The results also highlighted the importance of presence of active physicians at the time of discharge from the ICU. The higher mortality rate of patients who did not have their physicians present at the time of their discharge indicates that this factor is crucial and needs in-depth investigation. This effect was consistent between different genders and age groups. Results also indicated a higher mortality rate in teaching hospitals, which increased in 2015 compared with that in 2014. Discharge rates were observed to be higher during nighttime than during daytime. Comparing the difference between the distribution of deceased patients who were discharged “Alive” from the ICU but discharged “Expired” from the hospital in the presence or absence of active physicians at patient discharge shows that the absence of physicians at patient discharge from the ICU resulted in a higher mortality rate.

It is worth noting that although this database follows a standardized method for documenting the data, certain issues regarding data quality remain. The database contains significant amounts of missing values for certain variables. We have also observed some inconsistent data in patient diagnosis information, specifically regarding the status of diagnosis upon discharge. Moreover, some unusual values in patient demographic information such as height and weight have been observed throughout this study. Furthermore, a number of tables (e.g., vital sign measurements, care plan documentation, treatment information) of this large database, which could potentially bring more insights into this database, remained unexplored in the present study.

As the results of this study have demonstrated, there are different factors and determinants associated with patient mortality risk. Understanding the importance of these factors can lead to the design of more effective mechanisms and systems in hospitals to improve the outcomes of hospital stays for critical care patients. The importance of the patients’ disease, co-presence of diagnoses, and demographic factors in mortality rates suggest a need for improvements in the existing systems for assessing patient mortality risk scores. The results also underscored the importance of the quality of care and hospital characteristics in patient mortality rates, which warrant further research on hospital resource allocation and quality-of-care standards.

## Data Availability

All data is publicly available at https://eicu-crd.mit.edu/

## Footnotes

1 Patients who passed away

2 CCU: Coronary care unit, CTICU: Cardiothoracic ICU, CSICU: Quaternary cardiac surgical ICU, Med-surg ICU: Medical-surgical ICU, MICU: Medical ICU, Neuro ICU: Neurological ICU, SICU: Surgical ICU

## References

[1] Kelly, Fiona E., et al. “Intensive care medicine is 60 years old: The history and future of the intensive care unit.” Clinical medicine 14.4 (2014): 376–379.

[2] Adhikari, Neill KJ, et al. “Critical care and the global burden of critical illness in adults.” The Lancet 376.9749 (2010): 1339–1346.

[3] Celi, Leo Anthony, et al. ““Big data” in the intensive care unit. Closing the data loop.” American journal of respiratory and critical care medicine 187.11 (2013): 1157–1160.

[4] Johnson, Alistair EW, et al. “Machine learning and decision support in critical care.” Proceedings of the IEEE 104.2 (2016): 444–466.

[5] Bender, William, Cheryl A. Hiddleson, and Timothy G. Buchman. “Intensive Care Unit Telemedicine: Innovations and Limitations.” Critical care clinics 35.3 (2019): 497–509.

[6] United States, Congress, House. “Health insurance portability and accountability act of 1996.” Public law 104-191 (1996). Retrieved from http://www.eolusinc.com/pdf/hipaa.pdf

[7] Pollard, Tom J., et al. “The eICU Collaborative Research Database, a freely available multi-center database for critical care research.” Scientific data 5 (2018): 1–13.

[8] Shahin, Tala B., et al. “The Connected Intensive Care Unit Patient: Exploratory Analyses and Cohort Discovery From a Critical Care Telemedicine Database.” JMIR medical informatics 7.1 (2019): 1–12.

[9] Wagner, Clifford H. “Simpson’s paradox in real life.” The American Statistician 36.1 (1982): 46–48.

[10] Srinivasan, Karthik, et al. “Predicting high-cost patients at point of admission using network science.” IEEE Journal of Biomedical and Health Informatics 22.6 (2017): 1970–1977.

[11] Baek, Hyunyoung, et al. “Analysis of length of hospital stay using electronic health records: A statistical and data mining approach.” PloS one 13.4 (2018).

[12] Hunter, Alex, Leslie Johnson, and Alberto Coustasse. “Reduction of intensive care unit length of stay: The case of early mobilization.” The health care manager 33.2 (2014): 128–135

[13] Vollam, Sarah, et al. “Out-of-hours discharge from intensive care, in-hospital mortality and intensive care readmission rates: A systematic review and meta-analysis.” Intensive care medicine 44.7 (2018): 1115–1129.

[14] Yang, Si, et al. “Association between time of discharge from ICU and hospital mortality: A systematic review and meta-analysis.” Critical Care 20.1 (2016): 390.

[15] Abt, K. “Descriptive data analysis: A concept between confirmatory and exploratory data analysis.” Methods of information in medicine 26.2 (1987): 77–88.

[16] Behrens, John T. “Principles and procedures of exploratory data analysis.” Psychological Methods 2.2 (1997): 131–160.

[17] Ingeman, Annette, et al. “Quality of care and mortality among patients with stroke: A nationwide follow-up study.” Medical care (2008): 63–69.

